# Self-Reported Effects of IncobotulinumtoxinA on Headache with Migraine-like Characteristics in Participants with Traumatic Brain Injury vs. Anomalous Health Incidents Treated at a Single Specialty Center

**DOI:** 10.64898/2026.08.18.26360627

**Authors:** Ananya Tripathi, Jordan Llorin, David L. Brody

## Abstract

**Objective:** To describe the self-reported effects of incobotulinumtoxinA treatments on migraine-like headache in participants who experienced traumatic brain injury versus Anomalous Health Incidents.

**Background:** Persistent headache attributed to traumatic injury to the head has been widely recognized as among the most common sequelae of concussion/“mild” traumatic brain injury. Such persistent headaches often have migraine-like characteristics and are typically treated similarly to idiopathic migraine. Patients who have experienced Anomalous Health Incidents have also commonly reported migraine-like headaches, but to our knowledge, no reports describing treatment for persistent headaches attributed to Anomalous Health Incidents have been published.

**Methods:** We describe the self-reported effects of incobotulinumtoxinA treatments on headache with migraine-like characteristics in 19 participants with traumatic brain injury and 11 who had experienced Anomalous Health Incidents from a single center.

**Results:** Self-reported benefits from incobotulinumtoxinA treatments were generally similar and statistically indistinguishable between groups. The Headache Impact Test-6 score decreased by a mean of 12 points in the traumatic brain injury group and 9.5 points in the Anomalous Health Incidents group from baseline to peak efficacy (p = 0.43), with concomitant reductions in work/school hours lost (62% vs. 50%) and family/leisure hours lost (75% vs. 33%). Furthermore, reductions in headache frequency (67% for the traumatic brain injury group vs. 57% for the Anomalous Health Incidents group), headache severity (36% vs. 23%), headache duration (37% vs. 50%), nausea/vomiting (50% vs. 25%), photophobia (34% vs. 29%), phonophobia (30% vs. 37%), visual aura (50% vs. 29%), vestibular aura (50% vs. 33%), and other aura (21% vs. 25%) from baseline to peak efficacy were similar in both groups. Likewise, time from treatment to response (6.5 vs. 7 days), duration of response (10.2 vs. 9.1 weeks), adverse effects (3/19 for the traumatic brain injury group, 3/11 for the Anomalous Health Incidents group), and improved efficacy of concomitant abortive treatments (30% vs. 50% for pain, 50% vs. 55% for aura) did not differ between groups. Osmophobia and cogniphobia, when present, did not improve on average in either group. Notably, the mean duration of response was less than 12 weeks in both groups, with only 3 participants with traumatic brain injury and 1 participant who had experienced Anomalous Health Incidents reporting benefit beyond the typical 12-week incobotulinumtoxinA treatment interval.

**Conclusion:** Overall, these findings provisionally indicate that at least some patients who have experienced Anomalous Health Incidents may subjectively benefit from incobotulinumtoxinA treatment for persistent migraine-like headaches similarly to patients with traumatic brain injury. Limitations include the open-label, single-center, primarily retrospective design; small sample size; and limited representativeness. Further prospective controlled studies are needed to determine whether these groups truly respond similarly to incobotulinumtoxinA and other standard treatments.

**Plain Language Summary:** Persistent migraine-like headaches are common after traumatic brain injury and have also been reported after anomalous health incidents, but little is known about whether patients in these groups respond similarly to migraine-prevention treatment. In this single-center self-report study, participants with traumatic brain injury or anomalous health incidents described their symptoms before and after incobotulinumtoxinA treatment; both groups reported generally similar improvements in headache frequency, headache severity, headache impact, and time lost from activities. These preliminary findings suggest that symptom-based migraine prevention may help some patients in both groups, although larger prospective controlled studies are needed to confirm these results.

**Trial Registration:** ClinicalTrials.gov, NCT07267819.

## INTRODUCTION

Headaches are a common sequela of traumatic brain injury (TBI)^1–4^ and Anomalous Health Incidents (AHI).^5,6^ Managing post-traumatic headaches for patients with TBI involves a multidisciplinary approach including behavioral therapy, physical therapy, oral medications, injectable medications, and interventional procedures. Essentially, these headaches can be treated similarly to the primary headache disorder they resemble.^7^ This approach is partly pragmatic, as systematic review evidence indicates that high-quality, post-traumatic headache-specific pharmacologic treatment studies remain limited.^8^ Onabotulinum toxin A and other medications in the class of botulinum toxins have been particularly effective at reducing the overall frequency and duration of headaches in patients with TBIs^9,10^, as well as non-TBI-related migraines. Additional background on traumatic brain injury, Anomalous Health Incidents, migraine-like headache, and botulinum toxin treatment is provided in the Supplemental Introduction.

There is limited literature on managing post-AHI headache disorders, and AHI patients face unique diagnostic and clinical management challenges.^11,12^ Initial observations suggested that migraines in patients with AHI are clinically similar to those seen in patients with TBI, and recent experience indicates that using multidisciplinary approaches similar to those used for TBI-related headache disorders may be appropriate.^6^ Botulinum toxin treatment has therefore been a commonly used modality in these patient populations in the Military Health System. However, there has been no research of which we are aware on the effects of botulinum toxin injections in patients with TBI vs. AHI. The aims of this study were to quantify similarities and differences in effects of migraine management with a specific form of botulinum toxin, incobotulinumtoxinA that is commonly used in the Military Health System between participants with TBI vs. AHI. By comparing self-reported efficacy of incobotulinumtoxinA in both groups, the aim was to assess for meaningful differences in treatment response.

## METHODS

The study was designed to explore whether there were differences between subjective responses to incobotulinumtoxinA in participants with AHI vs. participants with TBI. Due to the lack of research on the effects of botulinum toxin injections in patients with TBI vs. AHI, we did not have a prespecified hypothesis about what these differences might be, nor any preliminary data about the magnitude of any differences that could be used for power calculations. As such, the sample sizes were based on convenience.

We designed a 3-part self-report survey to capture information from participants about their experiences with headaches and responses to incobotulinumtoxinA. The first part addressed status prior to starting incobotulinumtoxinA. The second part addressed status during the peak of incobotulinumtoxinA efficacy. The third part addressed status after they subjectively felt that the incobotulinumtoxinA effects had worn off. Most participants were already being treated with incobotulinumtoxinA and were asked to retrospectively recall their clinical status prior to incobotulinumtoxinA treatment. If they did not experience a peak of incobotulinumtoxinA efficacy, they were asked to recall their clinical status approximately 4 weeks after incobotulinumtoxinA treatment. For participants assessed prospectively, they were asked to complete the second survey 4 weeks after their incobotulinumtoxinA treatment. If they did not experience wearing off, they were asked to report their status prior to their next incobotulinumtoxinA treatment or at approximately 12 weeks after incobotulinumtoxinA treatment. Additional details regarding headache classification, survey administration, eligibility criteria, secondary and exploratory outcomes, missing data, and confidentiality procedures are provided in the Supplemental Methods.

The study protocol was reviewed and approved by the Uniformed Services University Human Research Protections Program (USUHS.2025-154) on June 6, 2025, as an exempt protocol not requiring Institutional Review Board review. All participants provided written informed consent prior to participation in the study. The research activities consisted only of completing the surveys. IncobotulinumtoxinA treatments and other medical interventions were provided as part of standard clinical care. All patients were treated with 155–175 units of incobotulinumtoxinA using the PREEMPT protocol^13^ by the same treating physicians. Treatments were performed every 6–12 weeks; patients were encouraged to schedule appointments when they felt the effects wearing off and usually could be accommodated for repeat treatments within 1–2 weeks.

The study was registered on ClinicalTrials.gov NCT07267819, submitted on 9/11/2025 and posted on 12/5/2025. The goal sample size was set at 60 based on an estimate of the potential number of participants available and constraints of research team effort availability. No prespecified power calculations were performed because no information was available regarding expected effect sizes.

The prespecified primary outcome measures were

1. Percent change in migraine headache frequency from baseline to peak effect (or 4 weeks post-treatment) and to wearing off (or 12 weeks post-treatment).
2. Percent change in migraine headache intensity from baseline to peak effect and to wearing off.
3. Percent change in headache impact test (HIT-6) score from baseline to peak effect and to wearing off.

Potential participants were identified based on medical record review by the treating physician and referred to the study coordinator. Potential participants were contacted in person after routine clinic visits, by telephone, and by email.

Statistical analyses were performed using GraphPad Prism 10.6.1. Dichotomous variables were analyzed using 2-sided Fisher’s exact tests. Continuous variables were assessed for normal distributions using scatter plots and Shapiro-Wilk tests. For normally distributed data, 2-sided Student t-tests or Welch’s t-test when variances differed were used. For non-normally distributed variables, 2-sided Mann-Whitney U tests were used. Sample size varied by outcome because participants could skip individual survey items; missing data were not imputed. No outliers were excluded. No correction for multiple comparisons was performed, since the analyses were intended as exploratory.

## RESULTS

### Characteristics of the participants

Enrollment began in August 2025 and concluded in February 2026. All participants were enrolled from a single specialty center within the Military Health System. There were 59 potential participants with TBI and 17 with AHI who were contacted, of which 27 and 15 signed informed consent and 19 and 11 provided data respectively (Supplemental Figure 1). Enrollment stopped because of constraints on research team availability prior to the prespecified maximum sample size of 60 total participants.

Demographics for the participants with TBI vs. AHI are presented in Table 1. There were no significant differences in age, sex, race, ethnicity, educational attainment, or year of most significant TBI or AHI. All participants with TBI were military service members, whereas most participants with AHI who reported affiliations were federal civilians (p = 0.0002, Fisher’s exact test). Body mass index was greater in participants with AHI (median 29, interquartile range [25-30]) vs. participants with TBI (median 24, [22-27]; p = 0.037, Mann-Whitney U test, Hodges-Lehman median difference = 3.0 BMI units). Military Occupational Specialty/job-title distributions differed between groups, with participants with TBI more commonly reporting intelligence or medical roles and participants with AHI more commonly reporting management/administrative or diplomatic roles (*p* = 0.0209, Fisher’s exact test). Of note, the National Capital Region has relatively few military service members in combat roles.

**Table 1.**
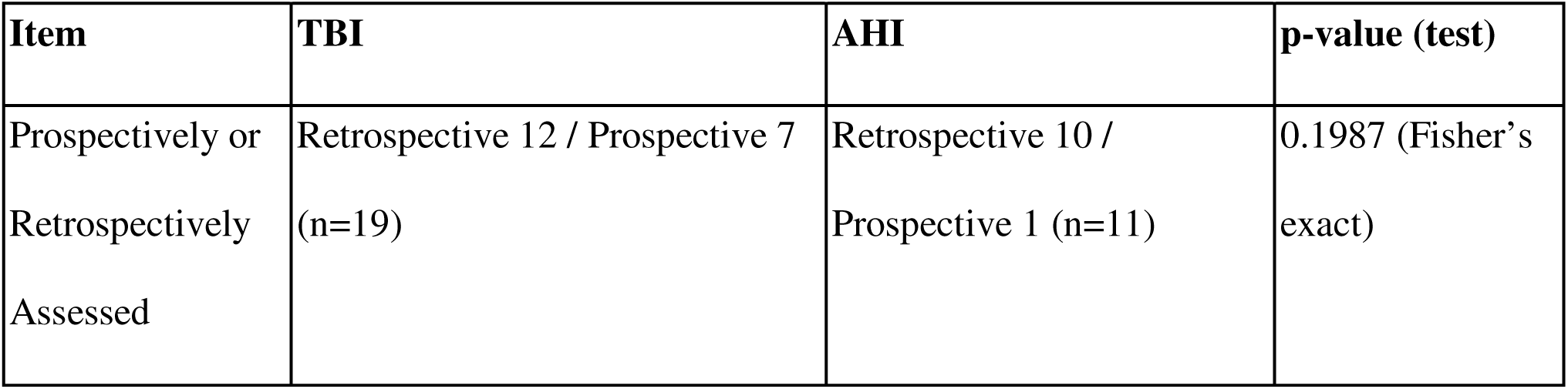
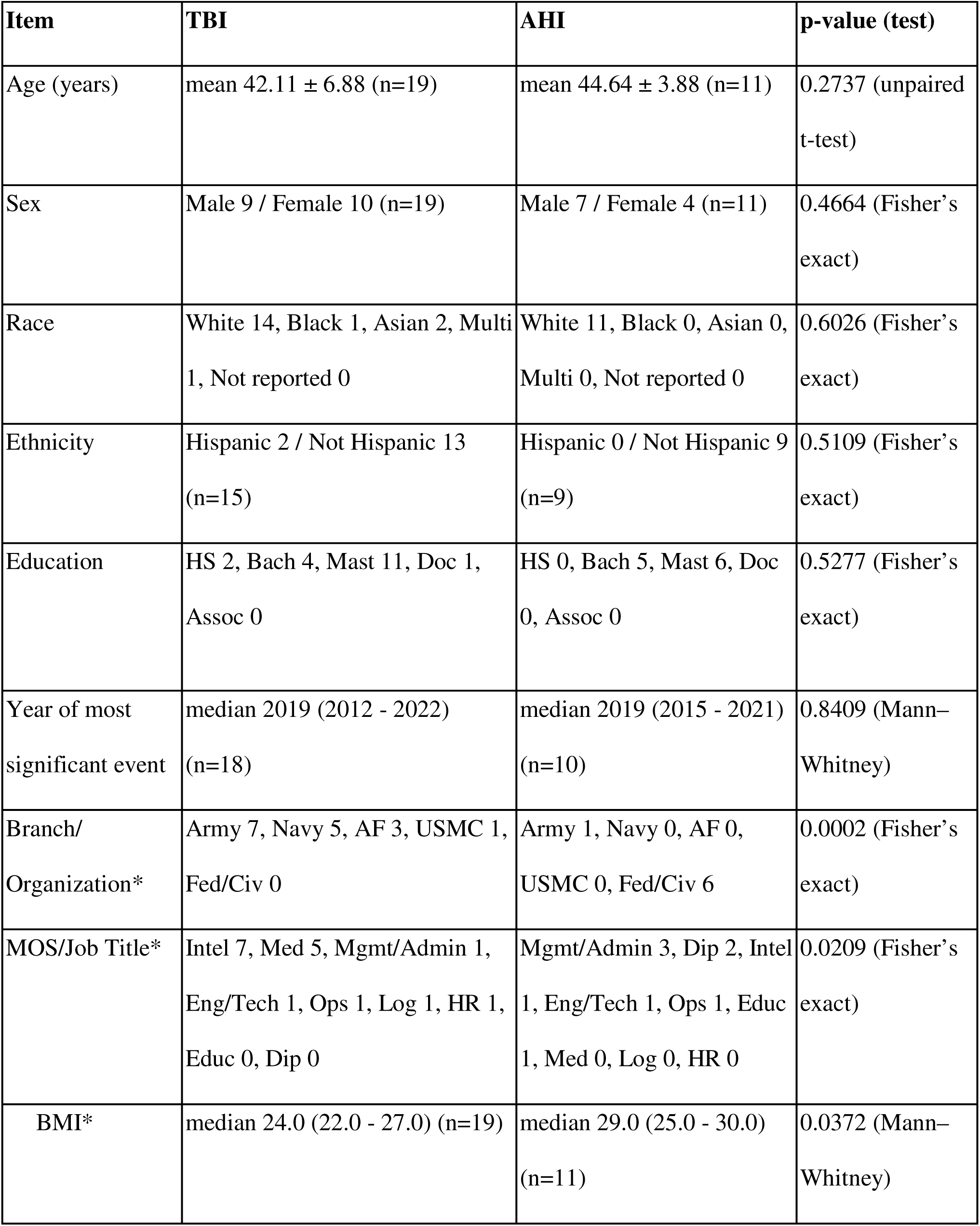
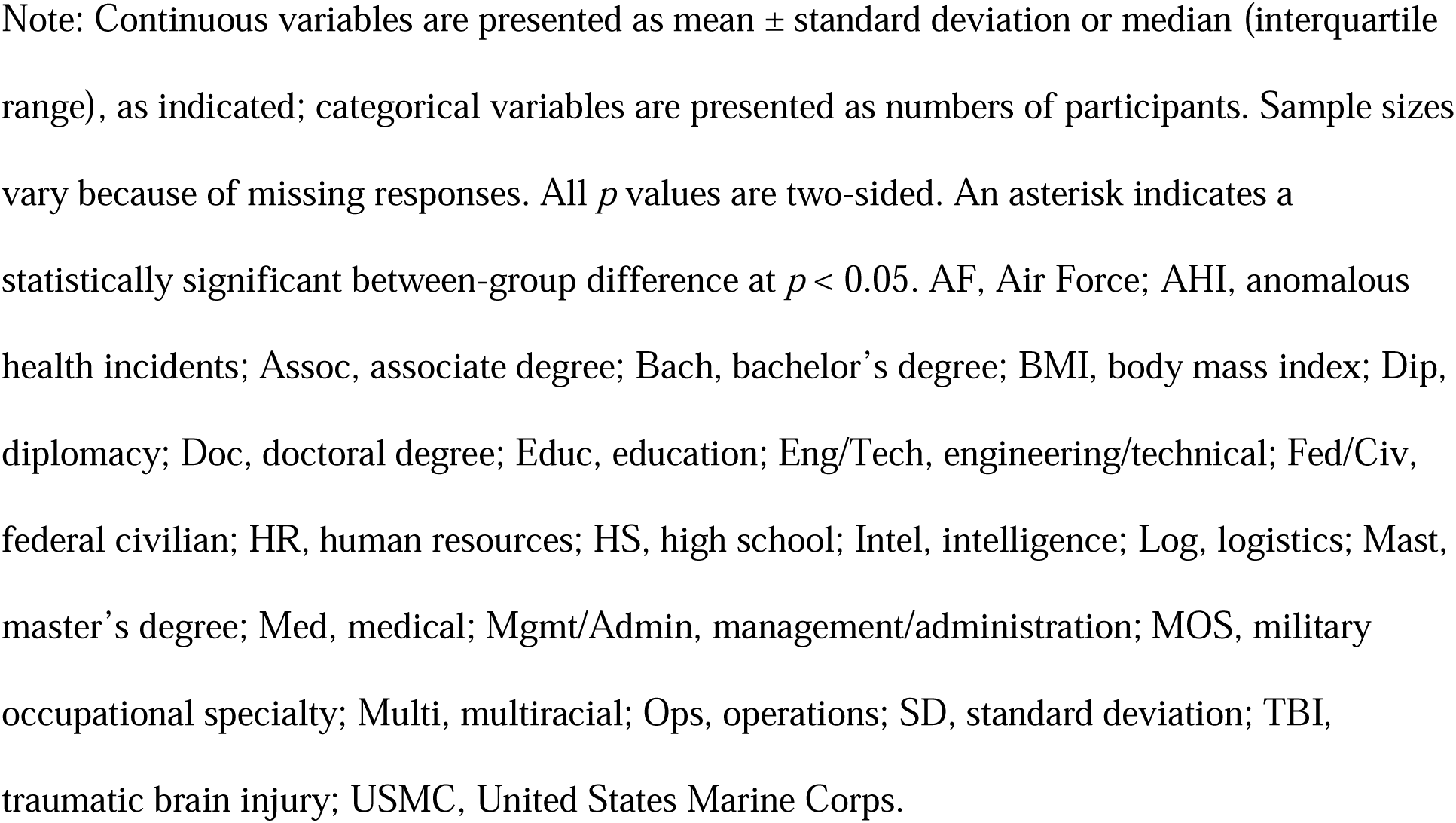
Participant demographic and clinical characteristics.

Self-reported lifestyle factors that may contribute to migraine outcomes are presented in Table 2. There were no significant differences in alcoholic drinks per week, maximum alcoholic drinks per 24 hours, self-reported sleep hours per night, and physical exercise hours per week. Most participants reported relatively healthy lifestyles.

**Table 2.**
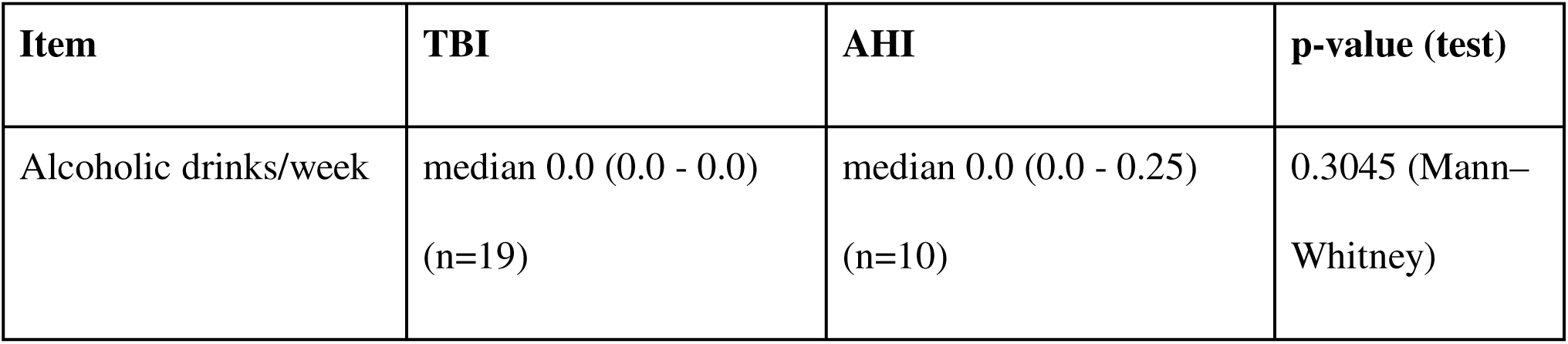
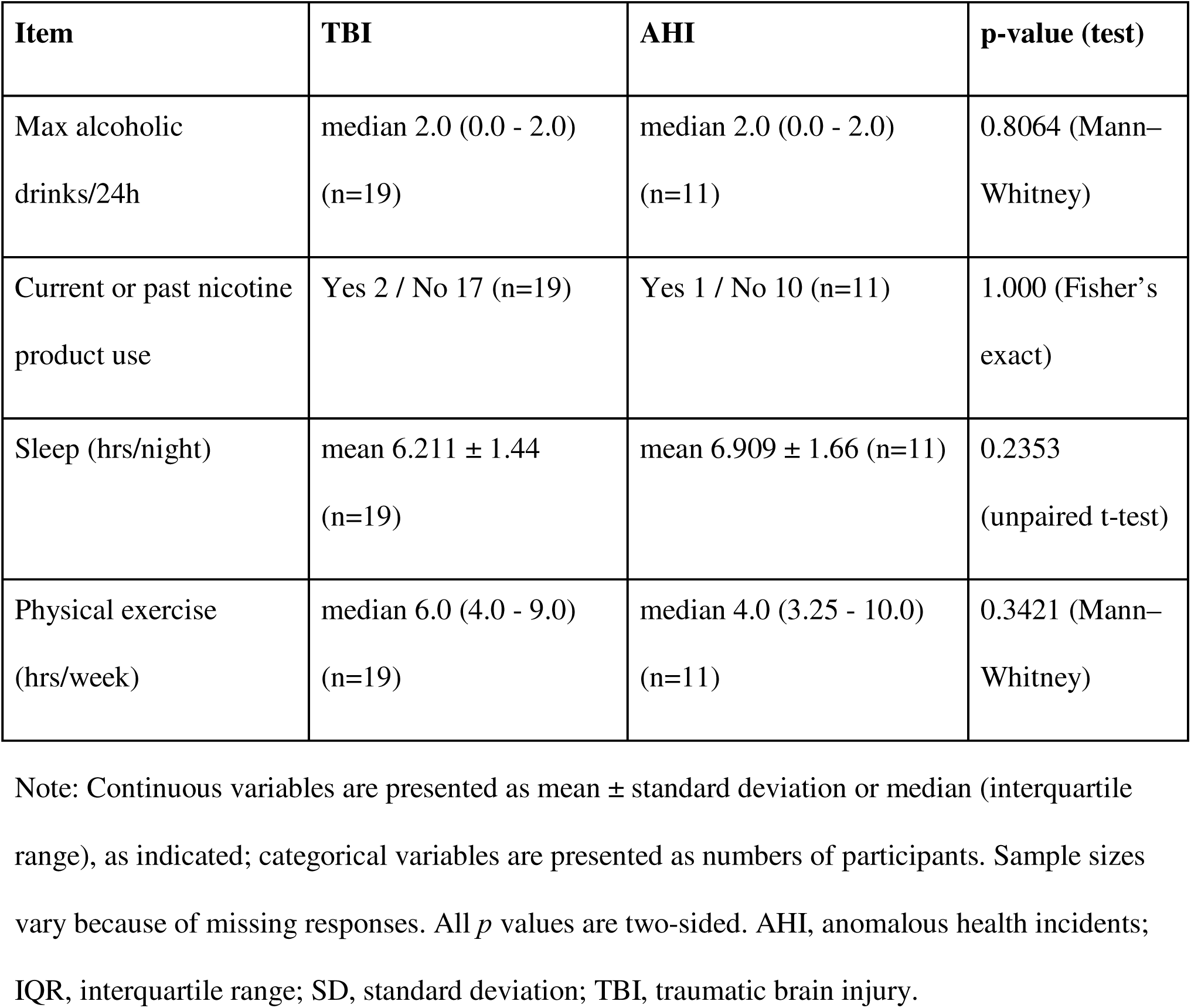
Self-reported lifestyle factors.

Self-reported comorbidities that may contribute to migraine outcomes are presented in Table 3. There were no significant differences in hypertension, diabetes, depression, PTSD, insomnia, obstructive sleep apnea, subjective cognitive impairment, tinnitus, hearing loss, or vestibular impairment. Anxiety was reported in a higher proportion (15/19) of participants with TBI vs AHI (4/11, p = 0.0465, Fisher’s exact test). Chronic pain was reported in a higher proportion (17/19) of participants with TBI vs. AHI (5/11, p = 0.0275). Vestibular impairment, which has been reported to be a hallmark of AHI, was reported in 9/11 participants with AHI and 8/19 with TBI (p = 0.0575). Participants in both groups had a high burden of comorbidities.

**Table 3.** Self-reported comorbidities that may contribute to migraine outcomes.

| Item | TBI | AHI | p-value (test) |
| --- | --- | --- | --- |
| Lifetime concussions*<br>(count) | median 3.0 (1.0 - 4.0)<br>(n=19) | median 0.0 (0.0 - 0.0) (n=11) | <0.0001 (Mann–Whitney) |
| Lifetime AHI events*<br>(count) | median 0.0 (0.0 - 0.0)<br>(n=16) | median 1.0 (1.0 - 2.0) (n=11) | <0.0001 (Mann–Whitney) |
| Hypertension | Yes 2 / No 17 (n=19) | Yes 2 / No 8 (n=10) | 0.5920 (Fisher’s exact) |
| Diabetes | Yes 0 / No 19 (n=19) | Yes 0 / No 11 (n=11) | >0.9999 (Fisher’s exact) |
| Depression | Yes 14 / No 5 (n=19) | Yes 4 / No 7 (n=11) | 0.0626 (Fisher’s exact) |
| PTSD | Yes 12 / No 7 (n=19) | Yes 3 / No 8 (n=11) | 0.1281 (Fisher’s exact) |
| Anxiety* | Yes 15 / No 4 (n=19) | Yes 4 / No 7 (n=11) | 0.0465 (Fisher’s exact) |
| Insomnia | Yes 9 / No 10 (n=19) | Yes 5 / No 6 (n=11) | >0.9999 (Fisher’s exact) |
| OSA | Yes 8 / No 11 (n=19) | Yes 2 / No 9 (n=11) | 0.2465 (Fisher’s exact) |
| Chronic pain* | Yes 17 / No 2 (n=19) | Yes 5 / No 6 (n=11) | 0.0275 (Fisher’s exact) |
| Subjective cognitive | Yes 9 / No 10 (n=19) | Yes 7 / No 3 (n=10) | 0.4335 (Fisher’s exact) |
| impairment |  |  |  |
| Tinnitus | Yes 17 / No 2 (n=19) | Yes 10 / No 1 (n=11) | >0.9999 (Fisher's exact) |
| Hearing loss | Yes 9 / No 10 (n=19) | Yes 6 / No 4 (n=10) | 0.6999 (Fisher's exact) |
| Vestibular impairment | Yes 8 / No 11 (n=19) | Yes 9 / No 2 (n=11) | 0.0575 (Fisher's exact) |

Note: Lifetime concussion and anomalous health incident event counts are presented as median (interquartile range); comorbidities are presented as numbers of participants reporting yes or no. Sample sizes vary because of missing responses. All variables were self-reported. All *p* values are two-sided. An asterisk indicates a statistically significant between-group difference at $p < 0.05$ . AHI, anomalous health incidents; IQR, interquartile range; OSA, obstructive sleep apnea; PTSD, posttraumatic stress disorder; TBI, traumatic brain injury.

Migraine history characteristics are presented in Table 4. Self-reported history of migraine prior to TBI or AHI was reported in a minority of participants. Similarly, self-reported family history of migraine was rare. Most participants were using or had used other treatments for migraine. These treatments included triptan, gepant, and other abortives; oral and injectable prophylactics other than incobotulinumtoxinA, and acupuncture. Approximately half had received other non-incobotulinumtoxinA botulinum toxin formulations.

**Table 4.** Migraine history characteristics.

| **Item** | **TBI** | **AHI** | **p-value (test)** |
| --- | --- | --- | --- |
| Treatment for migraine before the most significant TBI or AHI event | Yes 3 / No 15<br>(n=18) | Yes 3 / No 8<br>(n=11) | 0.6457 (Fisher's exact) |
| Family History of Migraine | Yes 3 / No 15 (n=18) | Yes 1 / No 10<br>(n=11) | >0.9999 (Fisher's exact) |
| Current migraine prophylactics | Yes 12 / No 7<br>(n=19) | Yes 6 / No 5<br>(n=11) | 0.7116 (Fisher's exact) |
| Current migraine abortives | Yes 15 / No 4<br>(n=19) | Yes 8 / No 3<br>(n=11) | 1.000 (Fisher's exact) |
| History of migraine prophylactics | Yes 9 / No 10<br>(n=19) | Yes 4 / No 7<br>(n=11) | 0.7084 (Fisher's exact) |
| History of migraine abortives | Yes 11 / No 8<br>(n=19) | Yes 6 / No 5<br>(n=11) | 1.000 (Fisher's exact) |
| Botulinum toxin formulations other than incobotulinumtoxinA | Yes 9 / No 6 (n=15) | Yes 4 / No 6<br>(n=10) | 0.4283 (Fisher's exact) |
| Other treatments for migraine, such as acupuncture or ketamine | Yes 9 / No 10<br>(n=19) | Yes 5 / No 6<br>(n=11) | 1.000 (Fisher's exact) |

| Item | TBI | AHI | p-value (test) |
| --- | --- | --- | --- |
| Year of first treatment with any botulinum toxin | median 2024<br>(2021.5–2025)<br>(n=19) | median 2022<br>(2022–2023)<br>(n=11) | 0.204 (Mann–Whitney) |

Note: Categorical variables are presented as numbers of participants reporting yes or no. Year of first treatment with any botulinum toxin is presented as median (interquartile range). Sample sizes vary because of missing responses. All *p* values are two-sided. AHI, anomalous health incidents; IQR, interquartile range; TBI, traumatic brain injury.

Self-reported baseline migraine characteristics prior to starting incobotulinumtoxinA were similar in participants with TBI vs. AHI (Figure 1). Migraine frequency was bimodal, with 7/11 participants with AHI and 5/19 participants with TBI reporting migraine every day (p = 0.06, Fisher’s exact test), while others reported 3-4 migraine days per week (Figure 1A). Baseline migraine frequency was not statistically greater in participants with AHI than TBI (p = 0.07, Mann-Whitney U test). Migraine duration was similarly bimodal, with many participants in both groups reporting 24 hours (continuous) migraine, and others reporting shorter duration migraines (Supplemental Figure 2). Pain intensity was high, with median of 8/10 in participants with TBI and 9/10 in participants with AHI (Figure 1B). Headache impact assessed using the HIT-6 indicated severe impact (>60) in nearly all participants (Figure 1C). Additional baseline migraine characteristics including migraine duration, photophobia, phonophobia, osmophobia, cogniphobia^14^, nausea/vomiting, visual aura, other aura, work hours lost, and leisure hours lost did not differ between groups (Supplemental Figure 2). Of note, vestibular aura was numerically more severe in participants with AHI vs. TBI (median 7/10 in AHI vs. 3/10 in TBI, Figure 1D) though the difference did not reach statistical significance. Visual and vestibular auras were reported in the majority of participants in both groups (Supplemental Figures 9 and 10). These results indicated that most participants in both groups reported characteristics similar to those of patients with severe chronic migraine with aura.

**Figure 1.**
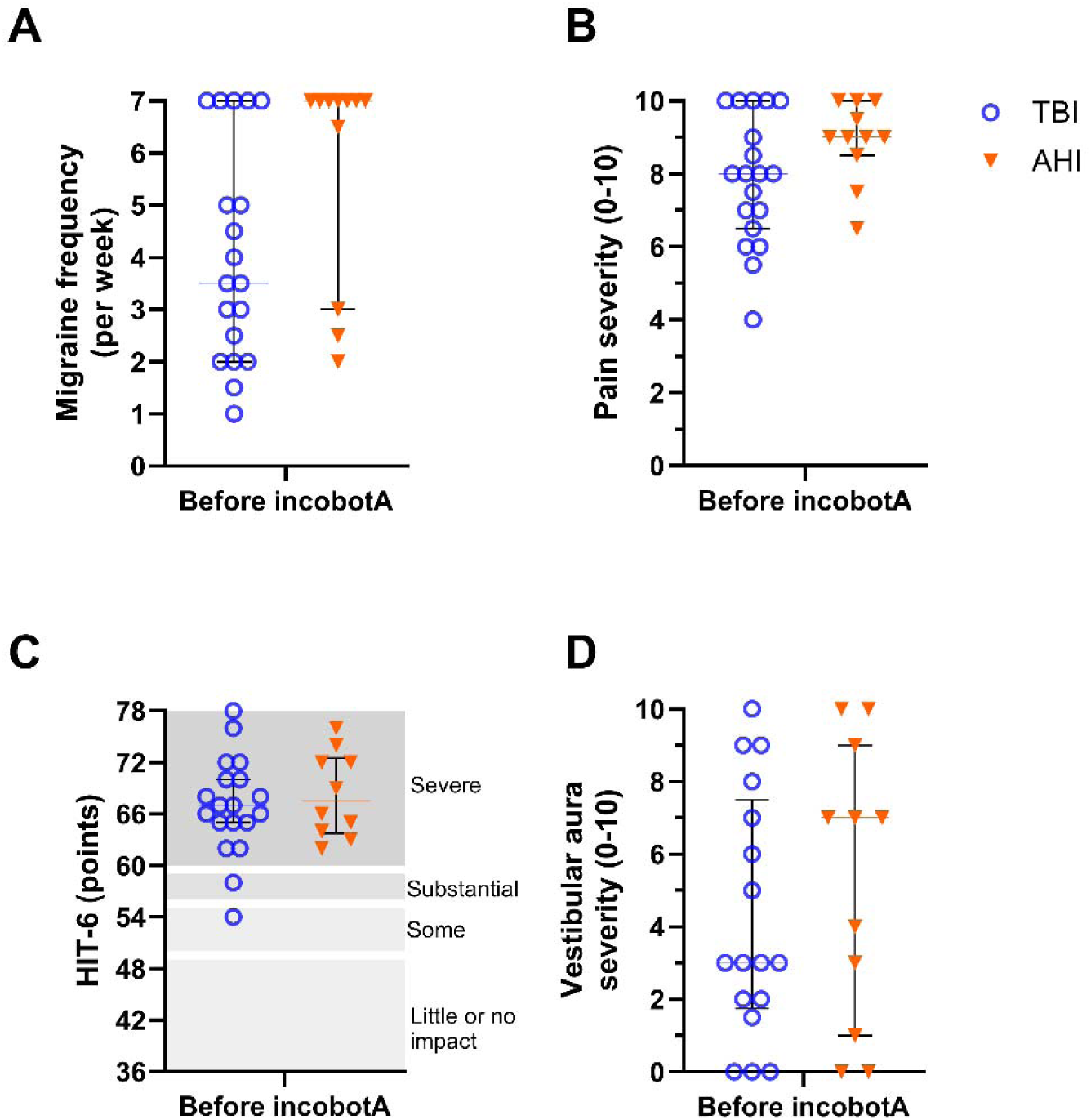
Baseline migraine characteristics before incobotulinumtoxinA treatment. Baseline migraine characteristics before incobotulinumtoxinA treatment (incobotA in the figure) are shown for participants with traumatic brain injury (TBI; blue circles) and anomalous health incidents (AHI; orange triangles). Panels show migraine frequency in days per week (A), pain severity rated from 0–10 (B), Headache Impact Test-6 (HIT-6) score (C), and vestibular aura severity rated from 0–10 (D). Individual points represent participants; horizontal bars indicate median and interquartile range. Migraine frequency was bimodal, with some participants reporting migraine every day and others reporting fewer migraine days per week. Pain severity was high in both groups, and HIT-6 scores indicated severe headache impact in nearly all participants. Vestibular aura severity was numerically higher in participants with AHI than TBI, but the difference did not reach statistical significance. Between-group comparisons were performed according to distribution, as described in the Methods; daily migraine frequency was also compared categorically using Fisher’s exact test.

### Self-reported effects of incobotulinumtoxinA treatments

Self-reported migraine characteristics were improved in both groups (Figure 2). Migraine frequency was the most substantially affected parameter in both groups (Figure 2A-C). Median relative reduction in self-reported migraine frequency from baseline to peak incobotulinumtoxinA efficacy was 67% in participants with TBI vs. 57% in participants with AHI (Figure 2A, p = 0.3 for comparison between groups). Even after participants reported that incobotulinumtoxinA effects had worn off, median reduction in self-reported migraine frequency from baseline was still 33% in participants with TBI vs. 38% in participants with AHI (p = 0.75 for comparison between groups). In absolute terms, median reduction in self-reported migraine frequency per week from baseline to peak incobotulinumtoxinA efficacy was 2 days in participants with TBI vs. 3 days in participants with AHI (Figure 2B, p = 0.94 for comparison between groups). After participants reported that incobotulinumtoxinA effects had worn off, median reduction in self-reported migraine frequency per week from baseline was still 1.25 days in participants with TBI vs. 1.5 days in participants with AHI (p = 0.94 for comparison between groups). While most participants reported a benefit, 2 participants with AHI reported no change in migraine frequency; individual trajectories are shown in Figure 2C.

**Figure 2.**
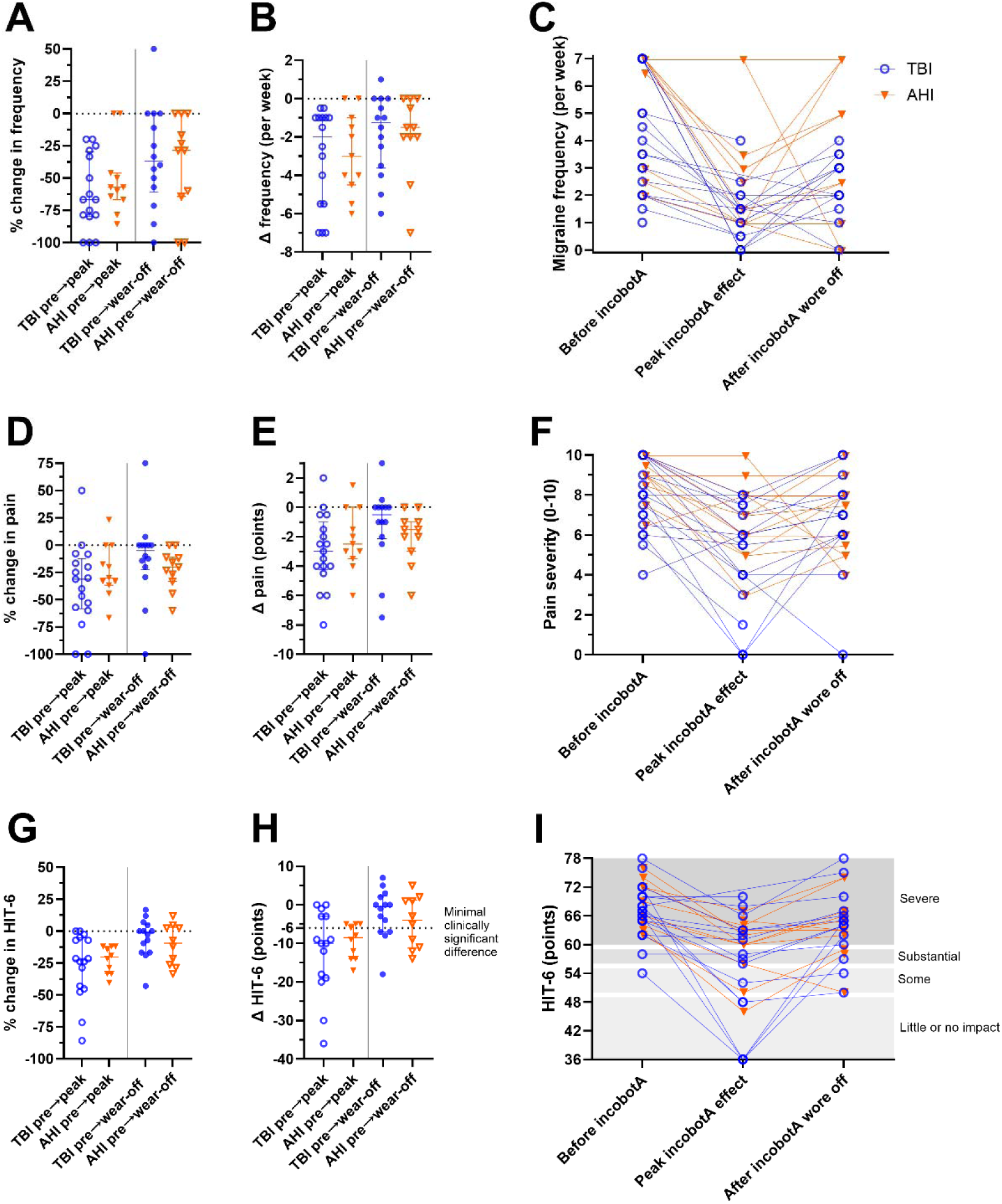
Self-reported effects of incobotulinumtoxinA on migraine frequency, pain severity, and headache impact. Self-reported migraine outcomes are shown before incobotulinumtoxinA treatment (incobotA in the figure), at peak incobotulinumtoxinA effect, and after participants reported that incobotulinumtoxinA effect had worn off for participants with traumatic brain injury (TBI; blue circles) and anomalous health incidents (AHI; orange triangles). Panels show percent change in migraine frequency (A), absolute change in migraine frequency in days per week (B), individual paired migraine frequency values across timepoints (C), percent change in pain severity (D), absolute change in pain severity on a 0–10 scale (E), individual paired pain severity values across timepoints (F), percent change in Headache Impact Test-6 (HIT-6) score (G), absolute change in HIT-6 score (H), and individual paired HIT-6 scores across timepoints (I). Individual points represent participants; horizontal bars indicate median and interquartile range for non-normally distributed variables and mean and standard deviation for approximately normally distributed variables. Dotted horizontal lines indicate no change from baseline. HIT-6 score interpretation bands are shown in panel I, and the dotted line in panel H indicates the minimal clinically important difference of 6 points. Migraine frequency, pain severity, and headache impact improved after incobotulinumtoxinA treatment in both groups, with no statistically significant between-group differences in change scores. Between-group comparisons of change scores were performed according to distribution, as described in the Methods section.

Self-reported migraine intensity was less dramatically reduced after incobotulinumtoxinA treatment in both groups (Figure 2D-F). Mean relative reduction in self-reported migraine intensity from baseline to peak incobotulinumtoxinA efficacy was 36% in participants with TBI vs. 23% in participants with AHI (Figure 2D, p = 0.32 for comparison between groups). After incobotulinumtoxinA effects had worn off, median reduction in migraine intensity from baseline was 5% in participants with TBI vs. 20% in participants with AHI (p = 0.14 for comparison between groups). In absolute terms, mean reduction in self-reported migraine intensity from baseline to peak incobotulinumtoxinA efficacy was 3 points on the 0–10 scale in participants with TBI vs. 2.2 points in participants with AHI (Figure 2E, p = 0.39 for comparison between groups). After incobotulinumtoxinA effects had worn off, median reduction in migraine intensity from baseline was still 0.5 points in participants with TBI vs. 1.5 points in participants with AHI (p = 0.14 for comparison between groups). While most participants reported a benefit, 3 participants reported no change, and 2 others reported increases in migraine intensity; individual trajectories are shown in Figure 2F.

Headache impact assessed using the HIT-6 was also substantially reduced after incobotulinumtoxinA treatment in both groups (Figure 2G-I). Mean relative reduction in HIT-6 scores from baseline to peak incobotulinumtoxinA efficacy were 27% in participants with TBI vs. 22% in participants with AHI (Figure 2G, p = 0.54 for comparison between groups). After incobotulinumtoxinA effects had worn off, mean reduction in HIT-6 scores from baseline were 5.3% in participants with TBI vs. 10.7% in participants with AHI (p = 0.41 for comparison between groups). In absolute terms, mean reduction in HIT-6 scores from baseline to peak incobotulinumtoxinA efficacy were 12 points in participants with TBI vs. 9.5 points in participants with AHI (Figure 2H, p = 0.43 for comparison between groups). A majority of participants with TBI (10/15) and AHI (6/10) improved more than the minimal clinically important difference of 6 points. After incobotulinumtoxinA effects had worn off, mean reduction in HIT-6 scores from baseline were 2.2 points in participants with TBI vs. 4.5 points in participants with AHI (p = 0.41 for comparison between groups). No participants reported worsening HIT-6 scores while 5 participants with TBI and 1 participant with AHI reported little to no headache impact after incobotulinumtoxinA treatment. Individual trajectories are shown in Figure 2I.

Self-reported migraine duration was also reduced after incobotulinumtoxinA treatment in both groups (Supplemental Figure 3). Median reduction in self-reported migraine duration from baseline to peak incobotulinumtoxinA efficacy was 37% in participants with TBI vs. 50% in participants with AHI (Supplemental Figure 3A, p = 0.67 for comparison between groups). After incobotulinumtoxinA effects had worn off, median self-reported migraine duration was back to baseline (median 0) in both groups (p = 0.21 for comparison between groups). In absolute terms, median reduction in self-reported migraine duration from baseline to peak incobotulinumtoxinA efficacy was 1.5 hours in participants with TBI vs. 1.0 hours in participants with AHI (Supplemental Figure 3B, p = 0.73 for comparison between groups). Many participants in both groups did not report any change in migraine duration, some even reported increased duration, and individual trajectories were variable (Supplemental Figure 3C).

Many associated migraine symptoms were reduced after incobotulinumtoxinA treatment in both groups, with no significant differences between groups. Nausea and vomiting were reduced 50% in the participants with TBI and 25% in the participants with AHI (Supplemental Figure 4). Photophobia was reduced 34% in the participants with TBI and 29% in the participants with AHI (Supplemental Figure 5). Phonophobia was reduced 30% in the participants with TBI and 37% in the participants with AHI (Supplemental Figure 6). Osmophobia (Supplemental Figure 7) and cogniphobia (Supplemental Figure 8) were less impacted.

Migraine auras diminished in severity after incobotulinumtoxinA treatment in both groups with no significant differences between groups. Most notably, visual auras were reduced in self-reported intensity by 50% in the participants with TBI and 29% in the participants with AHI (Supplemental Figure 9). Vestibular auras were reduced in self-reported intensity by 50% in the participants with TBI and 33% in the participants with AHI (Supplemental Figure 10). Other auras, when present, were also reduced (Supplemental Figure 11). In a few participants, auras were eliminated entirely.

These results indicated that most participants reported substantial improvements in migraine characteristics following incobotulinumtoxinA treatment, with no statistically significant differences between participants with AHI and TBI, and greatest magnitude of effects on migraine frequency.

### Timeline of self-reported incobotulinumtoxinA effects

Most participants reported that the benefits of incobotulinumtoxinA began within 14 days but did not last a full 12 weeks (Figure 3). There were no statistically significant differences between groups. Specifically, the median time to onset of incobotulinumtoxinA effects was 6.5 days in the participants with TBI and 7 days in the participants with AHI (Figure 3A, p = 0.69). The mean duration of benefit was 10.2 weeks in the participants with TBI and 9.1 weeks in the participants with AHI (Figure 3B, p = 0.57). Of note, only 3 participants with TBI and 1 participant with AHI reported that benefit lasted greater than 12 weeks, the typical frequency of incobotulinumtoxinA treatment.

**Figure 3.**
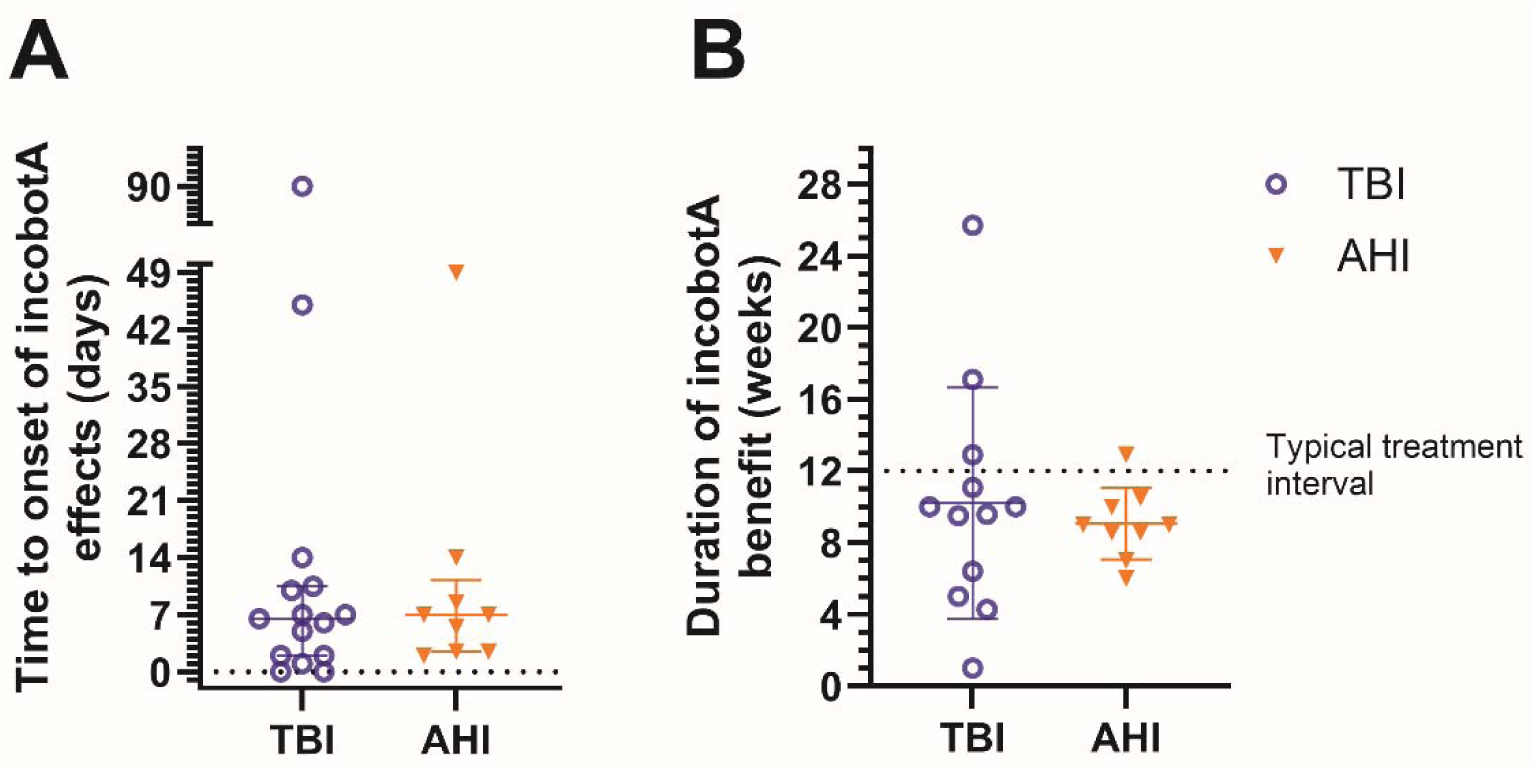
Timeline of self-reported incobotulinumtoxinA effects. Self-reported timing of incobotulinumtoxinA (incobotA in the figure) effects is shown for participants with traumatic brain injury (TBI; blue circles) and anomalous health incidents (AHI; orange triangles). Panel A shows time to onset of incobotulinumtoxinA effects in days. Panel B shows duration of incobotulinumtoxinA benefit in weeks. Individual points represent participants; horizontal bars indicate median and interquartile range in panel A and mean and standard deviation in panel B. Most participants reported onset of benefit within 14 days, and most reported that benefit lasted less than 12 weeks. The dotted horizontal line in panel B indicates the typical 12-week incobotulinumtoxinA treatment interval. Time to onset and duration of benefit did not significantly differ between groups. Between-group comparisons were performed using a two-tailed Mann–Whitney U test for time to onset and Welch’s unpaired t test for duration of benefit.

### Adverse effects

Most participants with TBI or AHI reported no adverse effects from incobotulinumtoxinA treatment (Supplemental Figure 12). Specifically, 1 participant with TBI and 1 with AHI reported an adverse effect characterized as severe, while 2 participants with TBI and 2 with AHI reported adverse effects characterized as mild (Supplemental Figure 12A). The adverse effects were all within the range of those expected for incobotulinumtoxinA treatment, including facial weakness, eye closure weakness, swelling, and persistent pain. None had serious adverse effects requiring hospitalization or discontinuation of incobotulinumtoxinA treatments. Duration of adverse effects ranged from 1 to 14 days (Supplemental Figure 12B).

### Use of acute abortive treatments for migraine

Participants reported their use of acute abortive medications before incobotulinumtoxinA treatment, 4 weeks after incobotulinumtoxinA treatment, and 12 weeks after incobotulinumtoxinA treatment. These data are presented descriptively in Supplemental Figures 13–15. No statistical analyses were performed, but there did not appear to be major differences between participants with AHI vs. TBI. The abortive medications in order from most frequent to least frequent were NSAIDs (with ibuprofen and Excedrin used more commonly than naproxen or indomethacin), triptans (with sumatriptan and rizatriptan used more commonly than zolmitriptan or eletriptan), acetaminophen, gepants (with ubrogepant used more commonly than rimegepant), and ergotamine (Supplemental Figure 13). After incobotulinumtoxinA treatment, many but not all participants in both groups reported that their abortive migraine medications were more effective than before incobotulinumtoxinA treatment in terms of pain relief, reduction in other symptoms, and reduction in aura (Supplemental Figure 14). Most participants reported no or mild side effects of abortive medications across timepoints, although some participants reported moderate or severe side effects (Supplemental Figure 15).

### Activity hours lost due to migraine

Participants in both groups reported reduced work and school hours lost due to migraine following incobotulinumtoxinA treatments (Figure 4A-C). Median reduction was 61.8% in the participants with TBI and 50% in the participants with AHI (Figure 4A). Median absolute reduction was 3 hours per week in the participants with TBI and 5 hours per week in the participants with AHI (Figure 4B). Most of the gains had dissipated by the time the incobotulinumtoxinA wore off. The individual trajectories are shown in Figure 4C. Likewise, participants in both groups reported reduced social, family, and leisure hours lost due to migraine following incobotulinumtoxinA treatments (Figure 4D-F). Median reduction was 75% in the participants with TBI and 33% in the participants with AHI (Figure 4D). Median absolute reduction was 5 hours per week in the participants with TBI and 4 hours per week in the participants with AHI (Figure 4E). Again, most of the gains had dissipated by the time the incobotulinumtoxinA wore off, especially in the participants with AHI. The individual trajectories are shown in Figure 4F.

**Figure 4.**
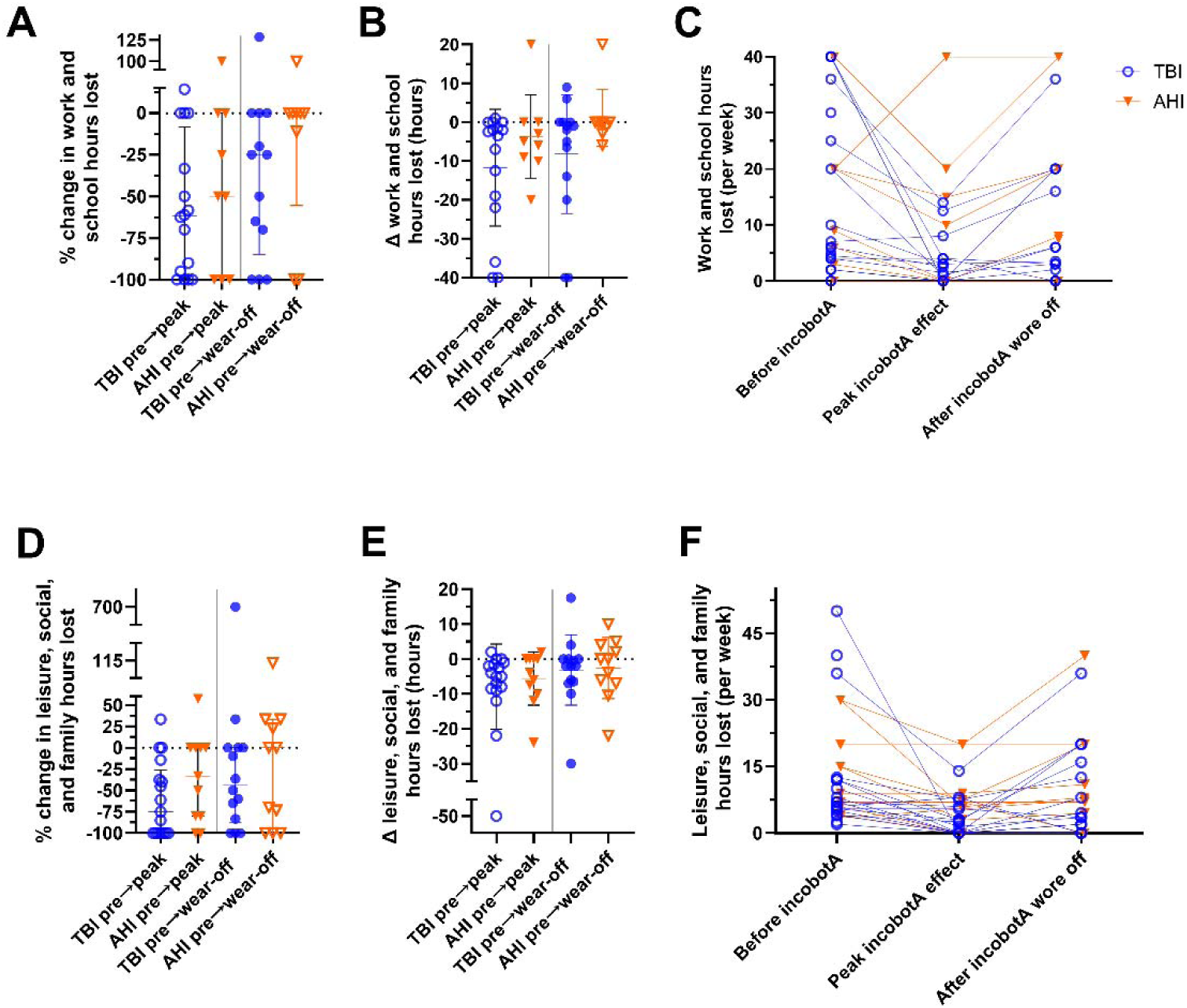
Self-reported activity hours lost due to migraine before and after incobotulinumtoxinA treatment. Self-reported activity hours lost due to migraine are shown before incobotulinumtoxinA treatment (incobotA in the figure), at peak incobotulinumtoxinA effect, and after participants reported that incobotulinumtoxinA effect had worn off for participants with traumatic brain injury (TBI; blue circles) and anomalous health incidents (AHI; orange triangles). Panels show percent change in work and school hours lost (A), absolute change in work and school hours lost in hours per week (B), individual paired work and school hours lost across timepoints (C), percent change in leisure, social, and family hours lost (D), absolute change in leisure, social, and family hours lost in hours per week (E), and individual paired leisure, social, and family hours lost across timepoints (F). Individual points represent participants; horizontal bars indicate median and interquartile range. Dotted horizontal lines indicate no change from baseline.

Participants in both groups generally reported fewer work/school and leisure/social/family hours lost at peak incobotulinumtoxinA effect, with most gains diminishing after participants reported that incobotulinumtoxinA effect had worn off. Between-group comparisons of change scores were performed according to distribution, as described in the Methods.

## DISCUSSION

In summary, participants with TBI and AHI reported similar improvements in migraine headache pain, migraine-related disability, other migraine symptoms, migraine auras, and lost activity time after incobotulinumtoxinA treatment. Time to onset and duration of benefit were also similar, with duration of benefit in both groups substantially shorter than the standard 12-week interval between treatments. Participants in both groups reported improved benefits from their other acute abortive medications after incobotulinumtoxinA treatment. IncobotulinumtoxinA treatments were well tolerated, with few adverse effects of which most were mild. Responses were generally similar to those previously reported in idiopathic migraine and other groups of patients with TBI. Additional discussion of clinical observations, mechanistic considerations, comparison with prior literature, and future research directions is provided in the Supplemental Discussion.

To our knowledge, this report is the first to describe treatment response for any medical condition related to AHI. Since headaches with migraine-like features are common, and incobotulinumtoxinA treatments are relatively standardized, this represented a logical initial investigation. Further strengths of this study include the standardized and comprehensive self-reported outcome measures, relatively uniform incobotulinumtoxinA treatments across all participants, and lack of apparent confounding by potential medicolegal biases. However, there were numerous limitations: 1) The study was open-label, primarily retrospective, and based at a single center. It was not randomized, blinded, or necessarily representative of the more general populations of patients with TBI and AHI. It is likely that many of the most severely affected patients were referred to our center, and that the characteristics of our cohort may represent an extreme portion of the TBI and AHI populations. 2) Because most participants completed their questionnaires retrospectively (n = 22) rather than prospectively (n = 8), the findings are subject to recall bias. 3) Only a portion of the patients treated at our center consented to participate, and not all of those who consented actually provided data. It is also likely that those who felt their incobotulinumtoxinA injections were effective were more likely to participate in the study than those who didn’t feel they benefited from them. This further limits the generalizability of the findings. In addition, the final sample size was smaller than the originally planned target of 60 participants, which limited statistical power and increased uncertainty around between-group comparisons. 4) In the absence of insurance-based constraints, incobotulinumtoxinA treatments were provided more frequently than every 12 weeks in many cases. Thus, effects may have been greater in magnitude than would have been observed with standard 12-week treatment intervals.

Of note, no additional adverse effects nor apparent development of tolerance to incobotulinumtoxinA treatment were observed with more frequent dosing. However, the study was not designed to compare standard treatment intervals with shorter treatment intervals. 5) The participants were also receiving care for other conditions attributed to TBI or AHI, and it is not known whether treatment with incobotulinumtoxinA in isolation would have similar effects. 6) The participants mainly came from the surrounding referral region, and there was a relative lack of racial and ethnic diversity. 7) Many portions of the self-report questionnaire have not been validated.

Importantly, similar self-reported responses to incobotulinumtoxinA in participants with AHI vs. TBI should not be interpreted as indicating that patients with AHI have had traumatic brain injuries. The pathophysiologies underlying AHI are still unknown.

The results of this study are generally concordant with previous studies reporting effects of botulinum toxin prophylactic treatments. To reiterate, there was no placebo control in our study. The implications of these results are modest, since the study was small and not controlled. Nonetheless, we take a cautiously optimistic view of the potential for treatment response for migraine symptoms and impact in patients with both TBI and AHI. In our view, the lack of pathophysiological understanding of AHI should not represent an impediment to empirical off-label symptom-based treatment using therapeutics designed for other indications. As more information is learned about AHI, it is possible that more specific, mechanism-based interventions may emerge. But in the interim, the Defense Health Agency recommendation is to treat symptoms and deficits using techniques drawn from TBI and other disorders.^11^

In conclusion, this small self-report-based study provides preliminary information about the similar efficacy of migraine prophylaxis with incobotulinumtoxinA in participants with TBI vs. AHI treated at a single center.

### Financial Support

This work received no project-specific funding. Salary support for Jordan Llorin was provided by the U.S. Army Medical Research and Development Command for the project entitled “Acute AHI Investigations.”

## Data Availability

All data produced in the present study are available upon reasonable request to the authors

## Abbreviations

AHI: anomalous health incidents
BMI: body mass index
CGRP: calcitonin gene-related peptide
DHA: Defense Health Agency
HIT-6: Headache Impact Test-6
ICHD: International Classification of Headache Disorders
incobotA: incobotulinumtoxinA
NSAIDs: nonsteroidal anti-inflammatory drugs
PREEMPT: Phase III Research Evaluating Migraine Prophylaxis Therapy
TBI: traumatic brain injury.

## Author Contributions

Ananya Tripathi: Conceptualization; Methodology; Project administration; Writing – review and editing.

Jordan Llorin: Investigation; Data curation; Formal analysis; Visualization; Writing – review and editing.

David L. Brody: Supervision; Project administration; Writing – original draft; Writing – review and editing.

## Data Availability

The data underlying this study are not publicly available due to participant confidentiality restrictions. The study protocol permits publication of de-identified and aggregate results and prohibits sharing information that could identify individual participants. De-identified aggregate results are provided in the manuscript and Supplemental Information.

## Acknowledgements

The authors would like to thank Jack Furman and Elena Bartels for helpful discussions.

## Disclaimer

The opinions and assertions expressed herein are those of the author(s) and do not reflect the official policy or position of the Uniformed Services University of the Health Sciences, Walter Reed National Military Medical Center, the Department of Defense/Department of War, or the Henry M. Jackson Foundation for the Advancement of Military Medicine.

