## Supplemental Information for "Self-Reported Effects of IncobotulinumtoxinA on Headache with Migraine-like Characteristics in Participants with Traumatic Brain Injury vs. Anomalous Health Incidents Treated at a Single Specialty Center"

TABLE OF CONTENTS

Supplemental Introduction	3

Supplemental Methods	6

Supplemental Results	9

Supplemental Figure 1. Participant flow diagram	9

Supplemental Figure 2. Other baseline migraine characteristics.	10

Supplemental Figure 3. Effects of incobotulinumtoxinA on self-reported migraine duration	11

Supplemental Figure 4. Effects of incobotulinumtoxinA on nausea and vomiting	12

Supplemental Figure 5. Effects of incobotulinumtoxinA on photophobia	13

Supplemental Figure 6. Effects of incobotulinumtoxinA on phonophobia	14

Supplemental Figure 7. Effects of incobotulinumtoxinA on osmophobia	15

Supplemental Figure 8. Effects of incobotulinumtoxinA on cogniphobia	16

Supplemental Figure 9. Effects of incobotulinumtoxinA on visual aura	17

Supplemental Figure 10. Effects of incobotulinumtoxinA on vestibular aura	18

Supplemental Figure 11. Effects of incobotulinumtoxinA on other aura	19

Supplemental Figure 12. Adverse effects of incobotulinumtoxinA treatments	20

Supplemental Figure 13. Migraine abortive medications reported	21

Supplemental Figure 14. Effectiveness of abortive medications	22

Supplemental Figure 15. Side effects of abortive medications	24

Supplemental Discussion	25

Supplemental References	28

**SUPPLEMENTAL INTRODUCTION**

In patients with TBI, direct biomechanical forces lead to cellular injury and neuroinflammation, contributing to post-traumatic headaches. Specifically, the disruption of trigeminovascular pathways and the release of inflammatory mediators, such as calcitonin gene-related peptide (CGRP), are implicated in the pathophysiology of post-traumatic migraines. Furthermore, the presence of comorbid conditions, such as sleep and mood disorders, can exacerbate headache symptoms and complicate treatment strategies.^1^ In TBI, imaging studies, post-mortem findings, and animal models have revealed important insights about the pathophysiology underlying headache disorders. However, the absence of clear structural damage on conventional neuroimaging in many AHI cases, and the lack of post-mortem studies or validated animal models raise questions about the underlying pathophysiological mechanisms.^2,3^ Theories involving unknown exposure and subsequent disruption of white matter function have been proposed. Some analyses suggest that the symptoms of AHI could result from the disruption of neuroimmune and neurotransmission mechanisms, similar to those affected in patients with TBI.^4,5^

In patients with TBI, post-traumatic migraine can be treated in the acute setting with triptans, NSAIDs, ergot alkaloids, and antiemetics.^6,7^ Chronic prophylactics can include tricyclic antidepressants, beta-blockers, CGRP-targeting agents and anti-seizure medications.^8,9^ Results of a recent clinical trial indicated that nightly prazosin reduced frequency of post-traumatic headaches.^10^ A low omega-6/high omega-3 diet also was shown to reduce post-traumatic headache frequency and intensity.^11^

Onabotulinum toxin A and other medications in the class of botulinum toxins have been particularly effective at reducing the overall frequency and duration of headaches in patients with TBIs^12,13^, as well as non-TBI-related migraines. Data from a mouse model of mild TBI-related headache behavior also indicate efficacy.^14^ Botulinum toxin treatment results in a blockade of neural stimulation by inhibiting the release of neurotransmitters at the presynaptic terminal.^15,16^ Botulinum toxin treatment is thought to reduce the exocytosis of pain mediators (substance P, CGRP, PACAP) from sensory neurons by cleaving SNAP25.^17,18^ This effect is temporary due to the formation of new synapses; thus, patients have to return for repeat injections every 10–12 weeks.

There is limited literature on managing post-AHI headache disorders, and AHI patients face unique diagnostic and clinical management challenges.^2,19^ Migraines can disrupt work, family, and leisure activities, and these challenges may be especially consequential in patients with AHI, for whom evidence-based treatment options remain limited.^19,20^

**Additional clinical context and rationale**

Recently, physicians from the participating specialty center within the Military Health System have been providing care for patients who have reported experiencing AHI. Care for Military Service Members with sequelae of AHI has been provided through the same facilities that provide care for Service Members with TBI. In addition, many non-military US Government employees with sequelae of AHI have been treated through the Secretarial Designee program. Care at military treatment facilities is not required; patients with AHI can also seek care through other healthcare systems.

By comparing self-reported efficacy of incobotulinumtoxinA in both groups at the participating specialty center, the aim was to assess for meaningful differences in treatment response. If detected, such differences could lead to more personalized and effective therapies that align with a holistic healthcare model in medicine. The findings could inform clinical guidelines and improve migraine management in patients with TBI and AHI, ultimately reducing the healthcare burden. This research was considered important for addressing the unmet needs of patients with AHI and advancing our understanding of treatment for headaches with migraine characteristics in diverse patient populations.

Several additional points should be noted about the clinical care for people with Anomalous Health Incidents (AHI) at the participating specialty clinical center.

1) Patients do not have to explain AHI to the providers or debate its existence; the providers at the participating center had all received standardized briefings about AHI.^21^ In contrast, patients have anecdotally reported that care in other contexts is complicated by lack of knowledge about AHI by providers, or disbelief in the existence of AHI. The position of the participating specialty center is that patients with AHI should receive medical care irrespective of as-yet-unresolved questions about causality or attribution.

2) Although many of the clinical approaches provided were developed for patients with TBI, none of the care provided is based on the assumption that the patients have had brain injuries. The care provided is symptom-based, not diagnosis or mechanism-based.

3) No one is asked to ‘prove’ that they have experienced an AHI, since at this time, there are no established objective diagnostic tests. In many respects, this is similar to care for patients with concussion/’mild’ TBI for which there are also no well-validated, objective diagnostic tests.

4) The military treatment facilities provide care for non-military personnel on a space-available basis; active-duty military service members typically have priority over non-military government employees. Nonetheless, regular care and frequent follow-ups are routinely provided for non-military government employees.

5) Because the Defense Health Program is independently self-funded, constraints placed by private health insurance or other government health programs do not necessarily apply. Treatment decisions at the participating specialty center occurred in the context of specialty clinical care within the Military Health System. As a result, treatment access and timing may not fully reflect constraints, authorization requirements, or scheduling intervals commonly encountered in other healthcare settings.

**SUPPLEMENTAL METHODS**

All participants in the study had headaches with migraine-like characteristics. We did not apply strict ICHD definitions of headache disorders. Some of the information required, such as when the headache began in relation to the TBI, was not available. Furthermore, there are no specific criteria for headaches related to AHI. The criteria required for headache attributed to other non-vascular intracranial disorder were difficult to apply since not enough is known about the underlying disorder (Criteria B) or typical headache characteristics (Criteria C3).^22^
Sample sizes varied by outcome because participants could skip individual survey items; missing data were not imputed. Percent change values were calculated only for participants with nonzero baseline values for the relevant outcome because percent change from a baseline value of zero is undefined. Adverse-effect and medication side-effect severity categories reflected participant self-report and were not independently adjudicated as formal clinical adverse-event severity grades. Data on acute abortive medication use are presented descriptively; medication dose, frequency of use, and number of uses per time interval were not systematically captured unless otherwise specified.

**Survey administration**

All participants completed the surveys themselves; no collateral source information was obtained. Participants were encouraged to consult their headache diaries if they kept them but were not required to do so. Surveys were completed on paper or in electronic format, in the clinic or at other locations. A research team member was available in person or by phone or email to answer questions about the survey (e.g. “What does cogniphobia mean?”). Completing the 3-part survey took about 20 minutes. No adverse effects of completing the surveys were reported. No compensation was provided. No biofluid sampling, imaging, or cognitive performance testing was performed for research.

**Eligibility criteria**

The inclusion criteria were:

- At least 18 years of age
- Able to provide written consent in English; no surrogate consent was allowed.
- An employee of the US Government, or an adult family member of a US Government employee
- Have received incobotulinumtoxinA treatment to prevent migraine related to TBI or AHI at a Military Treatment Facility or other US Medical Facility
- Able to participate in at least 80% of the assessments
- A US Citizen and not a dual national of the country where the research activities take place

**Secondary and additional outcome measures**

Prespecified secondary outcome measures were:

1) Time to onset of incobotulinumtoxinA effects
2) Duration of incobotulinumtoxinA benefits.

Additional, non-prespecified analyses included:

3) Headache duration
4) Intensity of nausea/vomiting
5) Intensity of photophobia, phonophobia, osmophobia, and cogniphobia
6) Intensity of visual aura, vestibular aura, and other aura
7) Frequency, duration, and severity of adverse effects
8) Changes in use and efficacy of concomitant abortive treatment for pain and aura
9) Changes in work, school, family, and leisure time missed due to migraine

**Data handling and confidentiality**

Paper records containing personally identifying information were transported in sealed envelopes and stored in a locked file cabinet. Electronic records were transmitted in encrypted format and stored in password-protected files on secure institutional computers. Participants had the option to include their individual data in their medical records and share it with their clinical providers but were not required to do so. The study protocol specified that there were no other data sharing or data use agreements in place. The protocol specified that only de-identified and aggregate results may be published and that individual data will not be shared in any way that would identify the participants.

**SUPPLEMENTAL RESULTS**

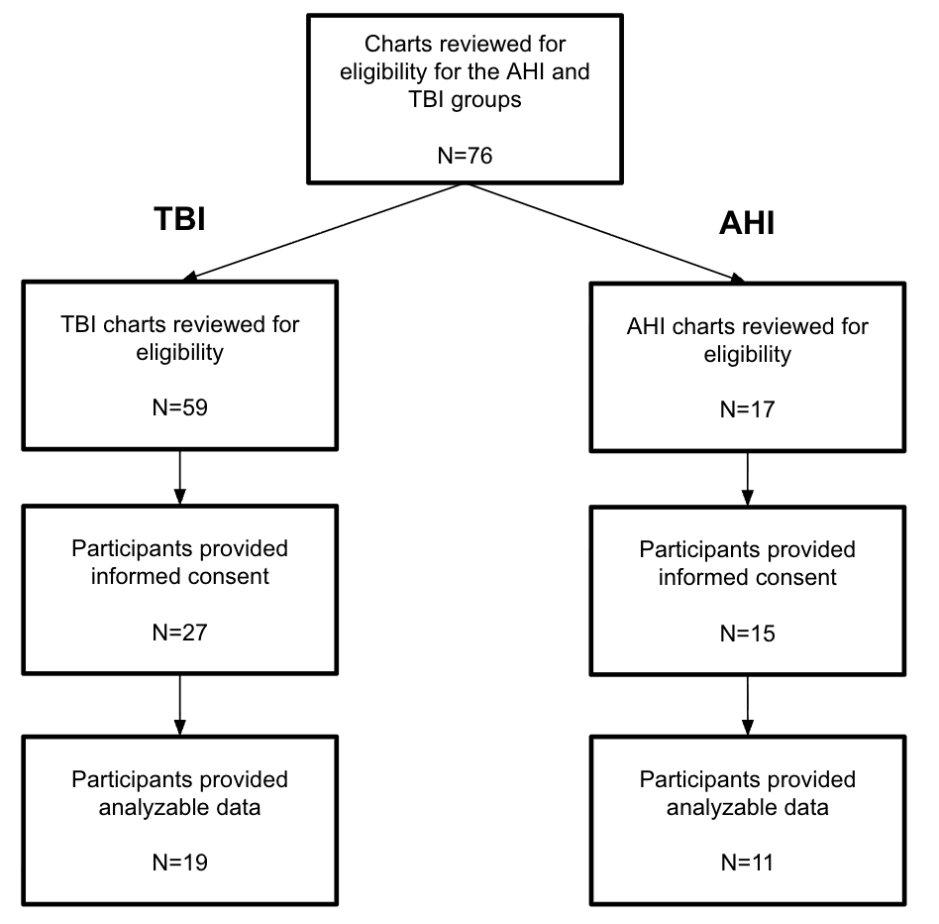

**Supplemental Figure 1. Participant flow diagram.**

Participant flow diagram showing chart review, informed consent, and inclusion in the traumatic brain injury (TBI) and anomalous health incidents (AHI) groups. A total of 76 charts were reviewed for eligibility, including 59 for the TBI group and 17 for the AHI group. Some individuals declined participation before providing informed consent. Among enrolled participants, 27 with TBI and 15 with AHI provided informed consent. Additional attrition occurred when some consented participants did not complete study survey/data collection or had missing or incomplete data. Data from 19 participants with TBI and 11 participants with AHI were ultimately included in the analyses.

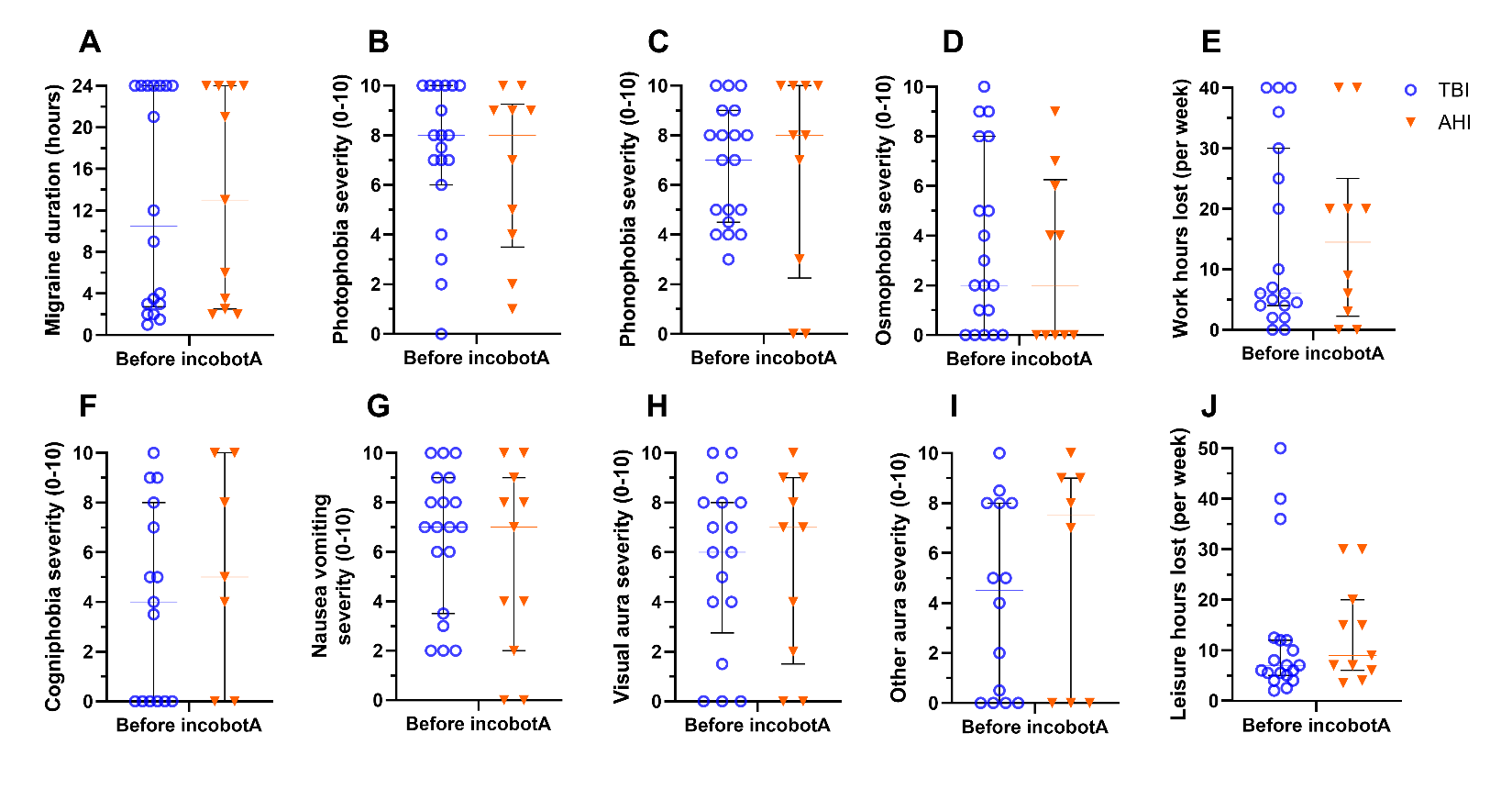

**Supplemental Figure 2. Other baseline migraine characteristics.**

Baseline migraine characteristics prior to incobotulinumtoxinA treatment (incobotA in the figure) are shown for participants with traumatic brain injury (TBI; blue circles) and anomalous health incidents (AHI; orange triangles). Panels show migraine duration (A), photophobia (B), phonophobia (C), osmophobia (D), work hours lost (E), cogniphobia (F), nausea/vomiting (G), visual aura (H), other aura (I), and leisure hours lost (J). Symptom severity ratings were reported on a 0–10 scale. Work and leisure hours lost were reported as hours per week. Individual points represent participants; horizontal bars indicate median and interquartile range. Migraine duration showed a similar pattern in both groups, with some participants reporting continuous 24-hour migraine and others reporting shorter-duration migraine. Exact two-tailed Mann–Whitney U tests showed no significant baseline differences between groups for any characteristic shown; all p ≥ 0.3884.

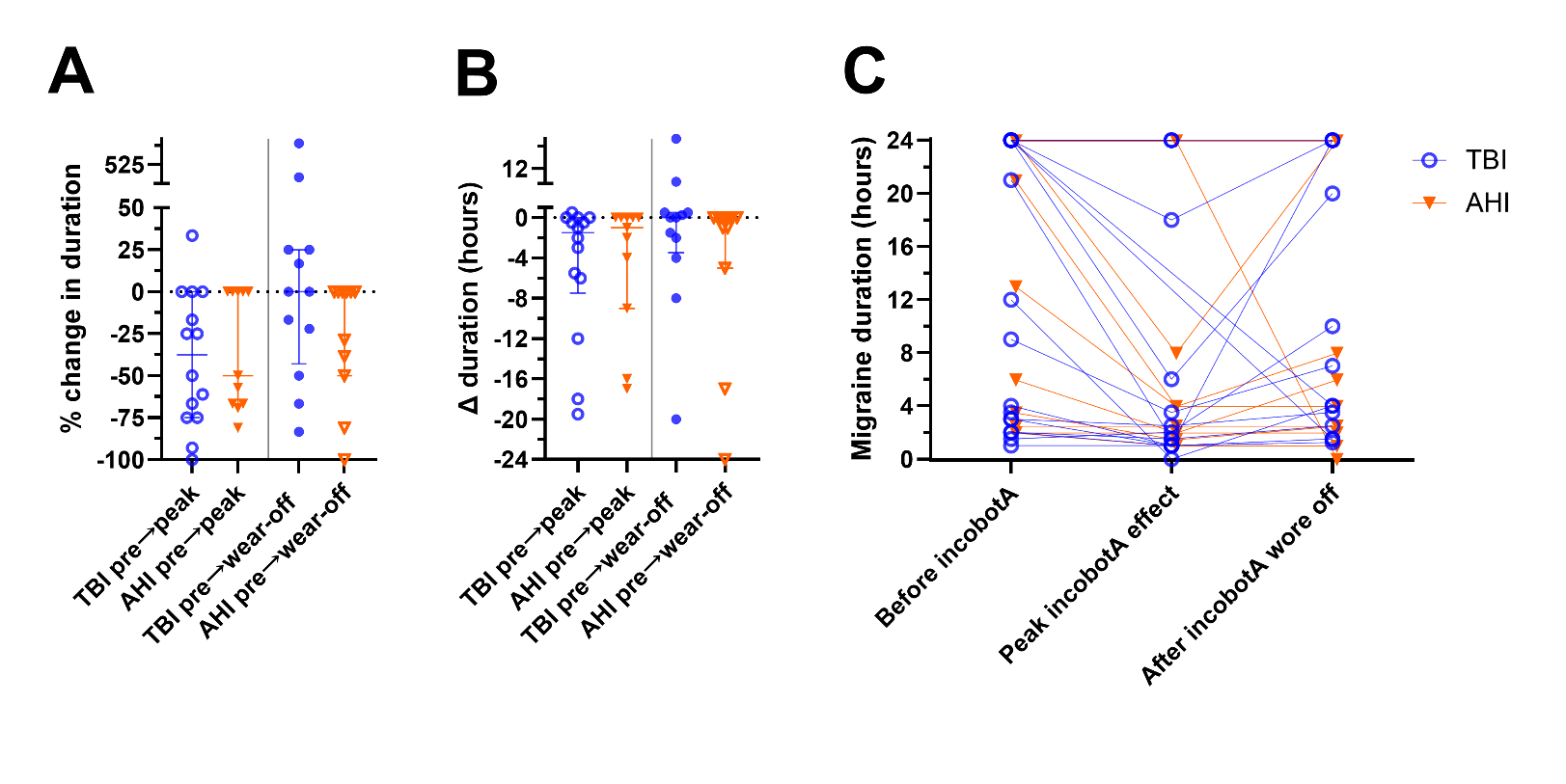

**Supplemental Figure 3. Effects of incobotulinumtoxinA on self-reported migraine duration.**

Self-reported migraine duration is shown before incobotulinumtoxinA treatment (incobotA in the figure), approximately 4 weeks after treatment when peak benefit was expected, and approximately 12 weeks after treatment when treatment effect had typically worn off for participants with traumatic brain injury (TBI; blue circles) and anomalous health incidents (AHI; orange triangles). Panels show percent change in migraine duration (A), absolute change in migraine duration in hours (B), and individual paired duration values across timepoints (C). Individual points represent participants; horizontal bars indicate median and interquartile range. Median migraine duration decreased from baseline to peak incobotulinumtoxinA effect in both groups, with a 37.5% reduction in TBI and 50.0% reduction in AHI, but the between-group difference was not significant (p = 0.671). In absolute terms, median reduction was 1.5 hours in TBI and 1.0 hour in AHI (p = 0.733). At approximately 12 weeks after treatment, median change from baseline was 0.0% and 0.0 hours in both groups (percent change, p = 0.208; absolute change, p = 0.299). Individual trajectories were variable, with many participants reporting no change and some reporting increased migraine duration. Between-group comparisons were performed using two-tailed Mann–Whitney U tests.

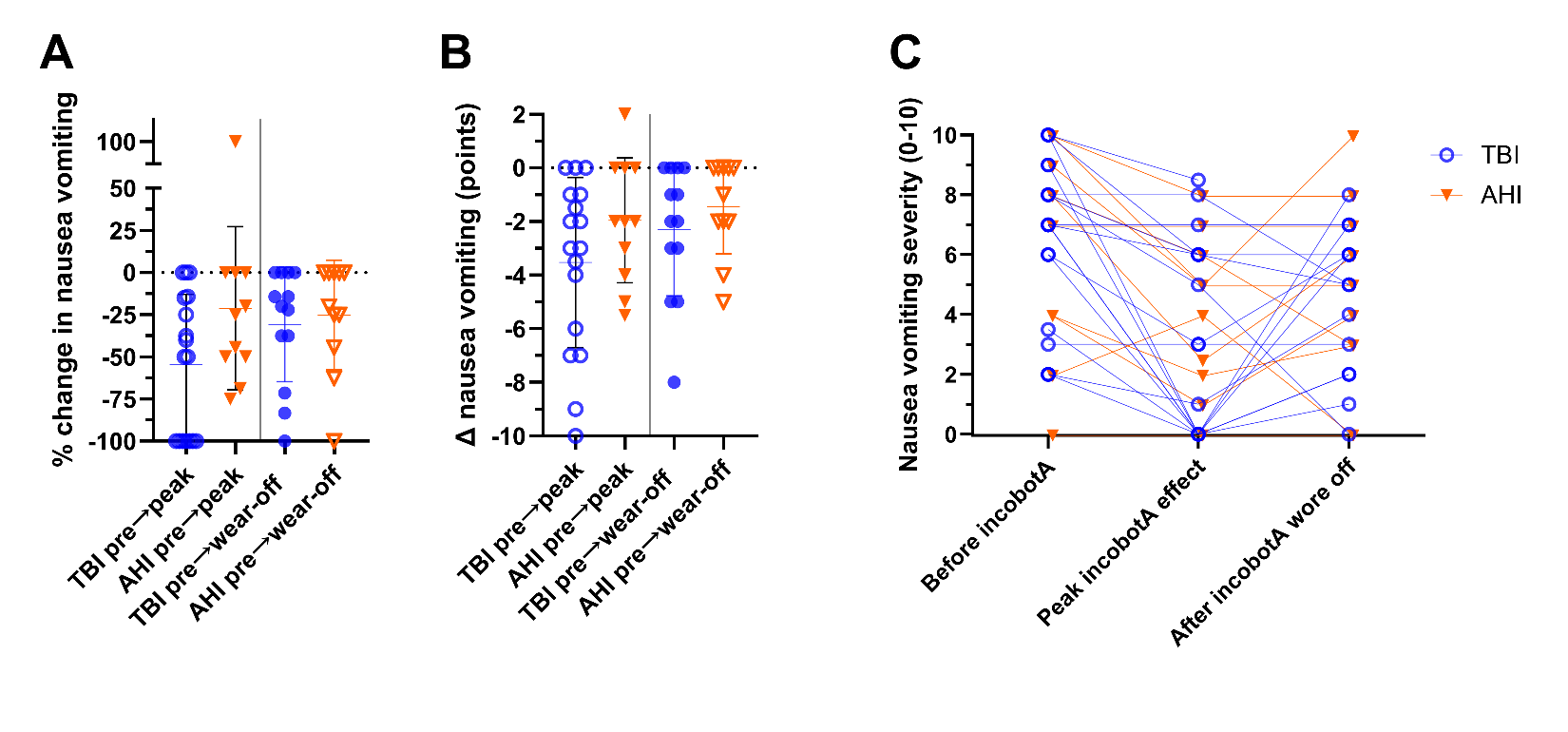

**Supplemental Figure 4. Effects of incobotulinumtoxinA on nausea and vomiting.**

Self-reported nausea/vomiting severity is shown before incobotulinumtoxinA treatment (incobotA in the figure), approximately 4 weeks after treatment when peak benefit was expected, and approximately 12 weeks after treatment when treatment effect had typically worn off for participants with traumatic brain injury (TBI; blue circles) and anomalous health incidents (AHI; orange triangles). Panels show percent change in nausea/vomiting severity (A), absolute change in nausea/vomiting severity (B), and individual paired severity ratings across timepoints (C). Nausea/vomiting severity was rated from 0–10. Individual points represent participants; horizontal bars indicate median and interquartile range. Nausea/vomiting severity decreased at peak treatment effect in both groups, with median reductions of 50.0% in TBI and 25.0% in AHI. Between-group comparisons of change scores were performed using two-tailed Mann–Whitney U tests, except for absolute change from baseline to peak treatment effect, which was analyzed using an unpaired t test. Percent change in nausea/vomiting severity did not significantly differ between groups from baseline to peak treatment effect (p = 0.125) or from baseline to approximately 12 weeks after treatment (p = 0.715). Absolute change also did not significantly differ between groups from baseline to peak treatment effect (p = 0.170) or from baseline to approximately 12 weeks after treatment (p = 0.400).

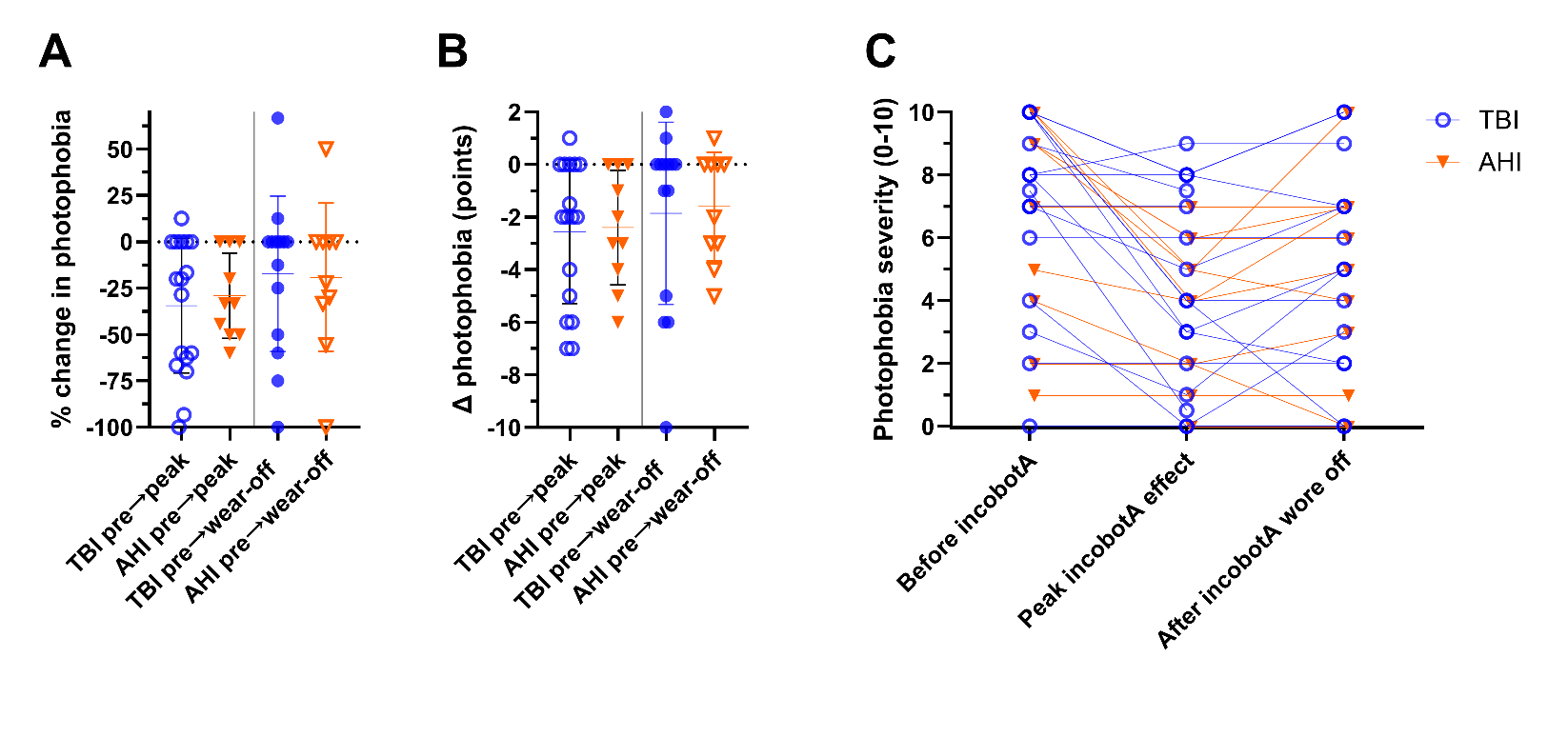

**Supplemental Figure 5. Effects of incobotulinumtoxinA on photophobia.**

Self-reported photophobia severity is shown before incobotulinumtoxinA treatment (incobotA in the figure), approximately 4 weeks after treatment when peak benefit was expected, and approximately 12 weeks after treatment when treatment effect had typically worn off for participants with traumatic brain injury (TBI; blue circles) and anomalous health incidents (AHI; orange triangles). Panels show percent change in photophobia severity (A), absolute change in photophobia severity (B), and individual paired severity ratings across timepoints (C). Photophobia severity was rated from 0–10. Individual points represent participants; horizontal bars indicate median and interquartile range. Photophobia severity decreased at peak treatment effect in both groups, with median reductions of 34.4% in TBI and 29.1% in AHI. Between-group comparisons of change scores were performed using two-tailed Mann–Whitney U tests, except for percent change from baseline to peak treatment effect, which was analyzed using an unpaired t test. Percent change in photophobia severity did not significantly differ between groups from baseline to peak treatment effect (p = 0.681) or from baseline to approximately 12 weeks after treatment (p = 0.800). Absolute change also did not significantly differ between groups from baseline to peak treatment effect (p = 0.992) or from baseline to approximately 12 weeks after treatment (p = 0.918).

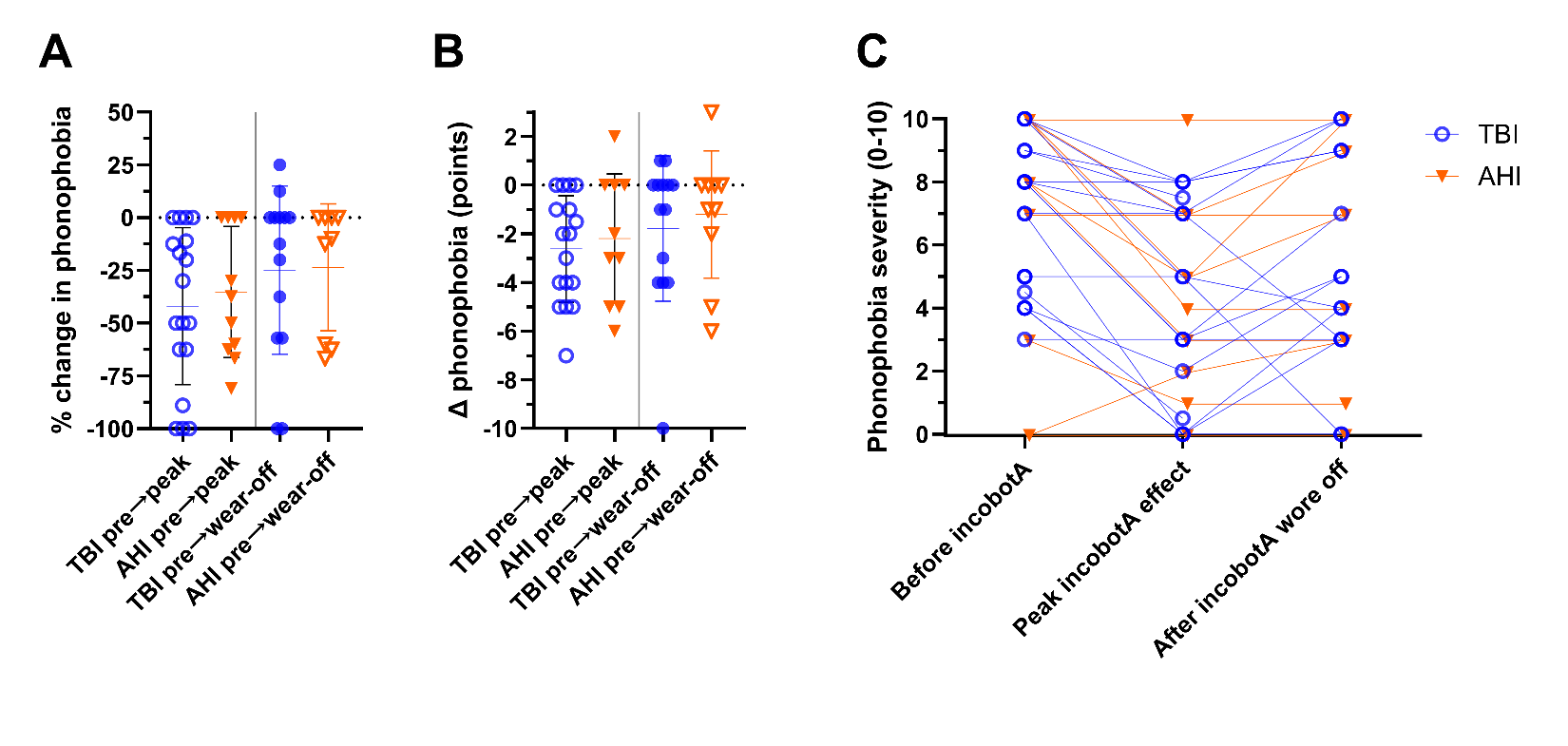

**Supplemental Figure 6. Effects of incobotulinumtoxinA on phonophobia.**

Self-reported phonophobia severity is shown before incobotulinumtoxinA treatment (incobotA in the figure), approximately 4 weeks after treatment when peak benefit was expected, and approximately 12 weeks after treatment when treatment effects had typically worn off for participants with traumatic brain injury (TBI; blue circles) and anomalous health incidents (AHI; orange triangles). Panels show percent change in phonophobia severity (A), absolute change in severity (B), and individual paired ratings across timepoints (C). Severity was rated from 0–10. Individual points represent participants; horizontal bars indicate median and interquartile range. Phonophobia severity decreased at peak treatment effect in both groups, with median reductions of 30.0% in TBI and 37.5% in AHI. Between-group comparisons were performed using two-tailed Mann–Whitney U tests, except for absolute change from baseline to peak treatment effect, which was analyzed using an unpaired t test. Neither percent nor absolute change differed significantly between groups from baseline to peak treatment effect (p = 0.718 and p = 0.662, respectively) or from baseline to approximately 12 weeks after treatment (p = 0.740 and p = 0.965, respectively).

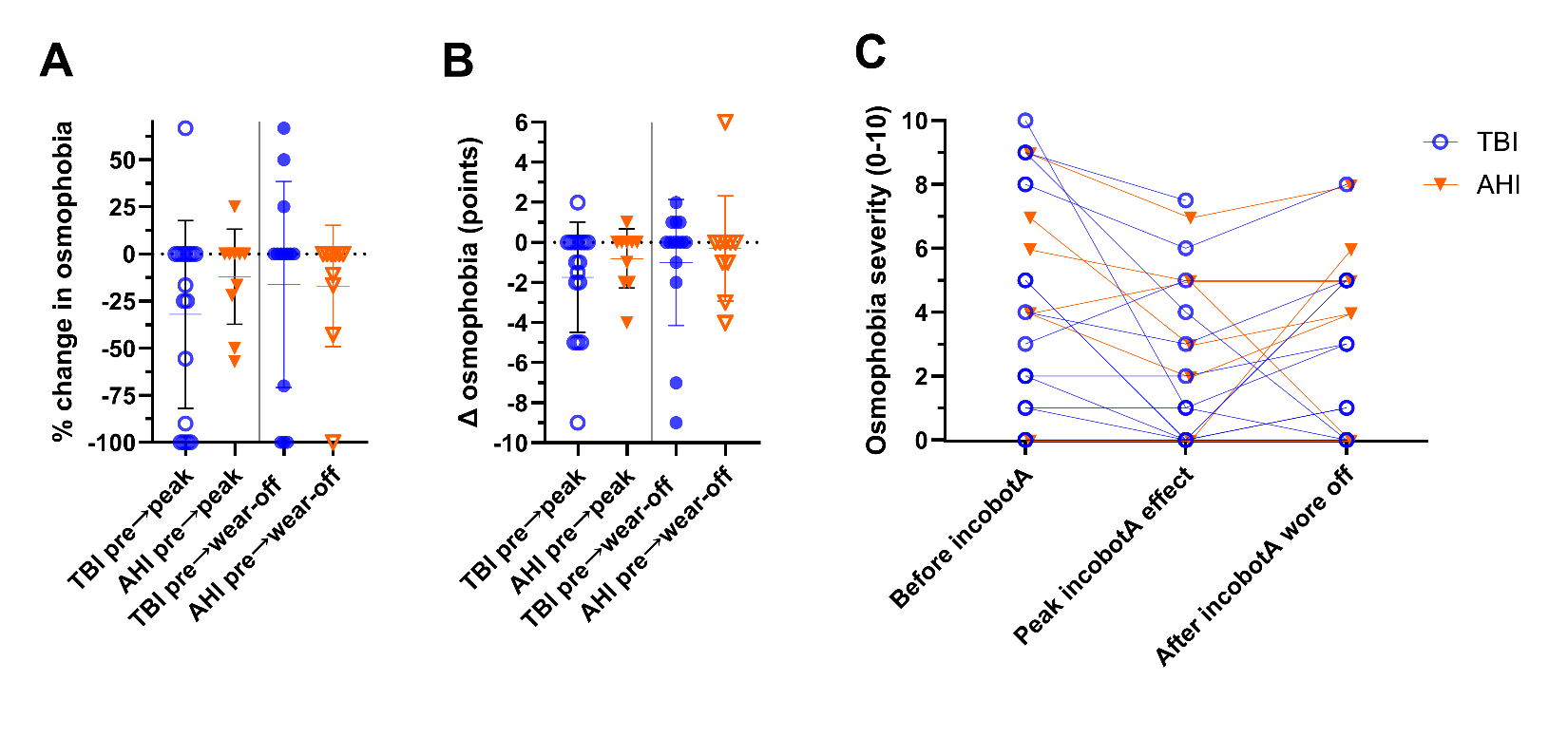

**Supplemental Figure 7. Effects of incobotulinumtoxinA on osmophobia.**

Self-reported osmophobia severity is shown before incobotulinumtoxinA treatment (incobotA in the figure), approximately 4 weeks after treatment when peak benefit was expected, and approximately 12 weeks after treatment when treatment effect had typically worn off for participants with traumatic brain injury (TBI; blue circles) and anomalous health incidents (AHI; orange triangles). Panels show percent change in osmophobia severity (A), absolute change in osmophobia severity (B), and individual paired severity ratings across timepoints (C). Osmophobia severity was rated from 0–10. Individual points represent participants; horizontal bars indicate median and interquartile range. Changes in osmophobia severity were modest and variable across participants. Between-group comparisons of change scores were performed using two-tailed Mann–Whitney U tests. Percent change in osmophobia severity did not significantly differ between groups from baseline to peak treatment effect (TBI median −16.7%, n = 17; AHI median 0.0%, n = 10; p = 0.280) or from baseline to approximately 12 weeks after treatment (TBI median 0.0%, n = 14; AHI median 0.0%, n = 10; p = 0.486). Absolute change also did not significantly differ between groups from baseline to peak treatment effect (p = 0.426) or from baseline to approximately 12 weeks after treatment (p = 0.566).

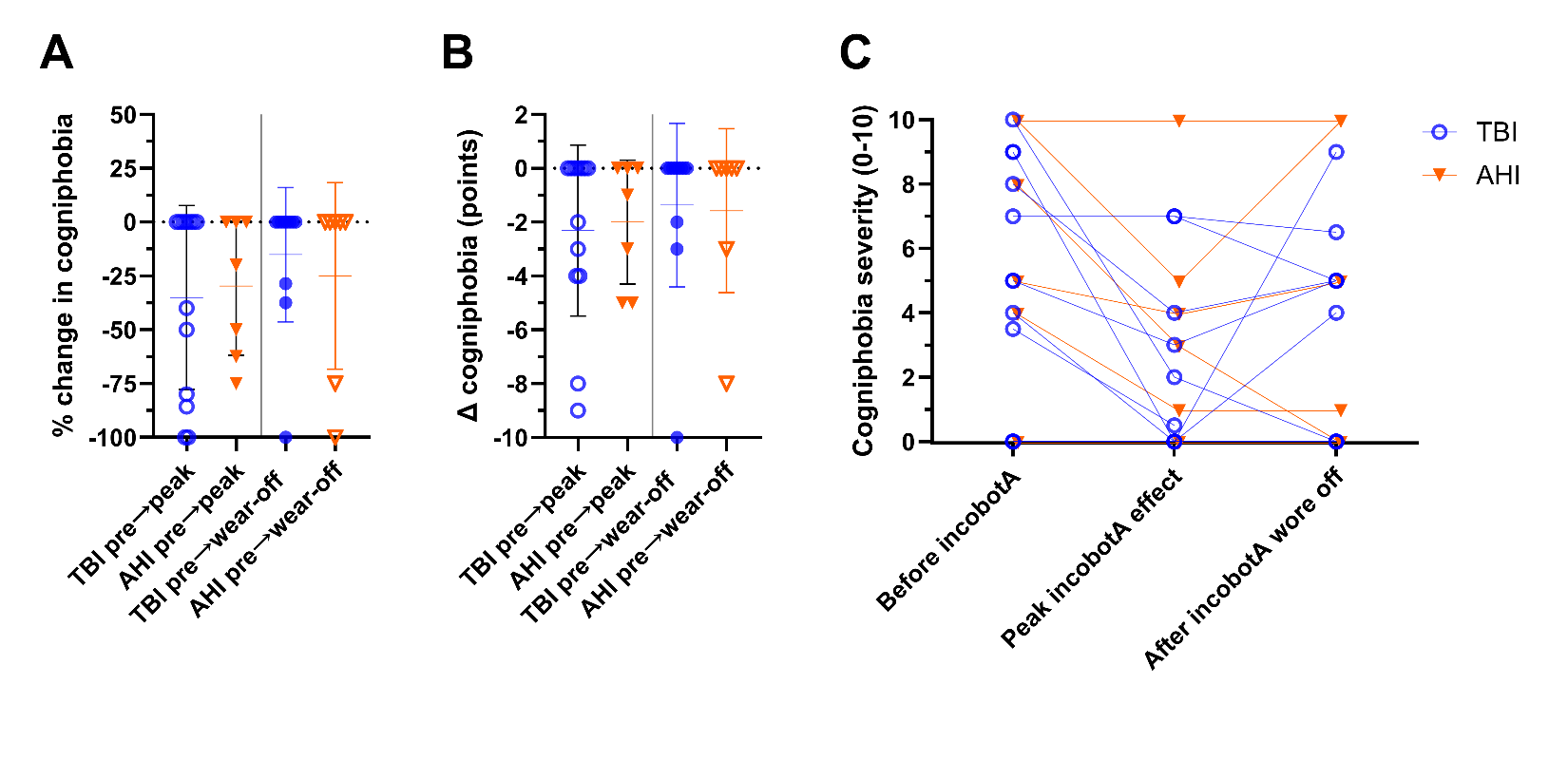

**Supplemental Figure 8. Effects of incobotulinumtoxinA on cogniphobia.**

Self-reported cogniphobia severity is shown before incobotulinumtoxinA treatment (incobotA in the figure), approximately 4 weeks after treatment when peak benefit was expected, and approximately 12 weeks after treatment when treatment effect had typically worn off for participants with traumatic brain injury (TBI; blue circles) and anomalous health incidents (AHI; orange triangles). Panels show percent change in cogniphobia severity (A), absolute change in cogniphobia severity (B), and individual paired severity ratings across timepoints (C). Cogniphobia severity was rated from 0–10. Individual points represent participants; horizontal bars indicate median and interquartile range. Changes in cogniphobia severity were modest and variable across participants. Between-group comparisons of change scores were performed using two-tailed Mann–Whitney U tests. Percent change in cogniphobia severity did not significantly differ between groups from baseline to peak treatment effect (TBI median 0.0%, n = 13; AHI median −1.0%, n = 7; p = 0.916) or from baseline to approximately 12 weeks after treatment (TBI median 0.0%, n = 11; AHI median 0.0%, n = 7; p = 0.757). Absolute change also did not significantly differ between groups from baseline to peak treatment effect (p = 0.858) or from baseline to approximately 12 weeks after treatment (p = 0.879).

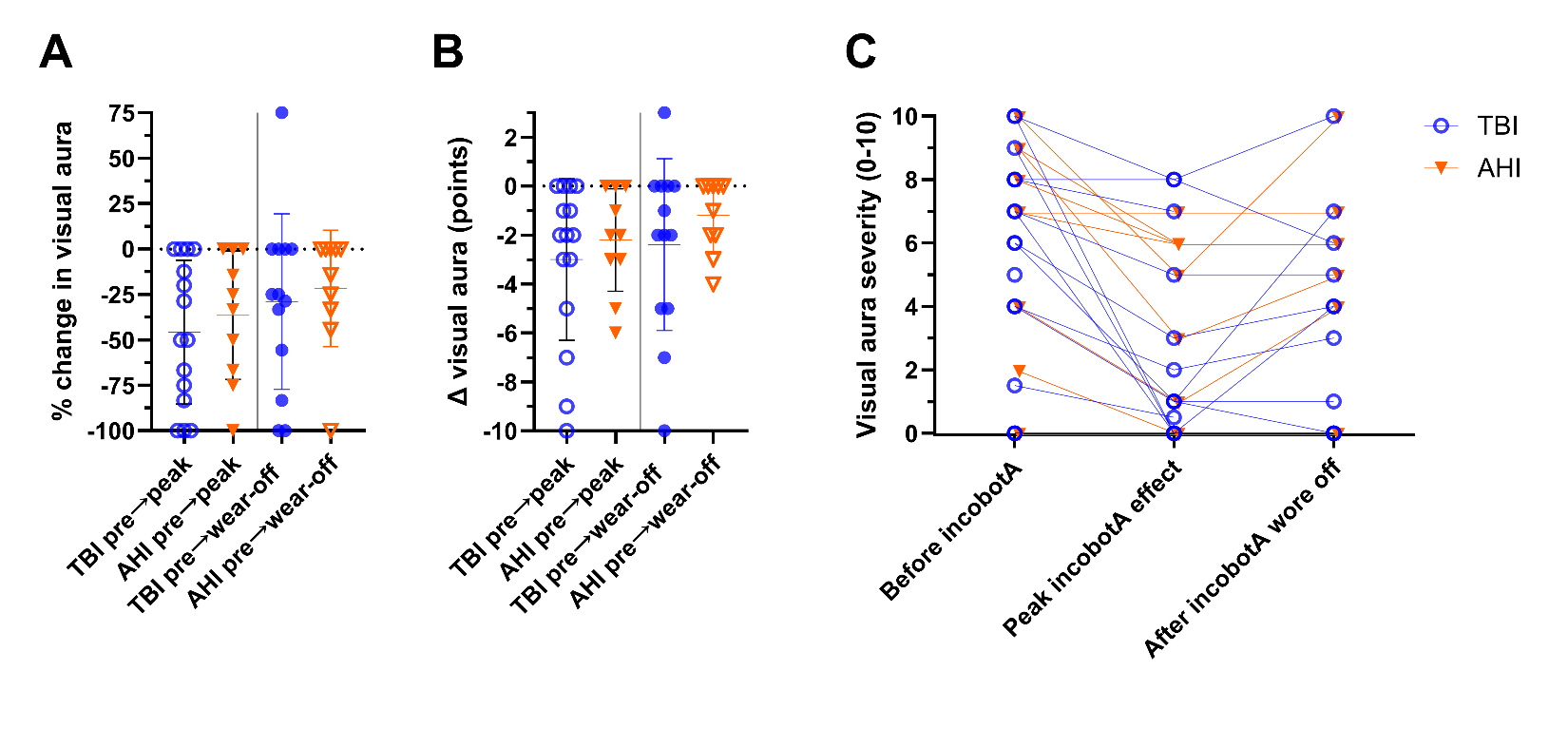

**Supplemental Figure 9. Effects of incobotulinumtoxinA on visual aura.**

Self-reported visual aura severity is shown before incobotulinumtoxinA treatment (incobotA in the figure), approximately 4 weeks after treatment when peak benefit was expected, and approximately 12 weeks after treatment when treatment effect had typically worn off for participants with traumatic brain injury (TBI; blue circles) and anomalous health incidents (AHI; orange triangles). Panels show percent change in visual aura severity (A), absolute change in visual aura severity (B), and individual paired severity ratings across timepoints (C). Visual aura severity was rated from 0–10. Individual points represent participants; horizontal bars indicate median and interquartile range. Visual aura severity decreased at peak treatment effect in both groups, with median reductions of 50.0% in TBI and 29.15% in AHI, but the between-group difference was not significant (p = 0.609). Percent change in visual aura severity did not significantly differ between groups from baseline to approximately 12 weeks after treatment (TBI median −25.0%, n = 13; AHI median −7.15%, n = 10; p = 0.556). Absolute change also did not significantly differ between groups from baseline to peak treatment effect (p = 0.772) or from baseline to approximately 12 weeks after treatment (p = 0.475). Between-group comparisons were performed using two-tailed Mann–Whitney U tests.

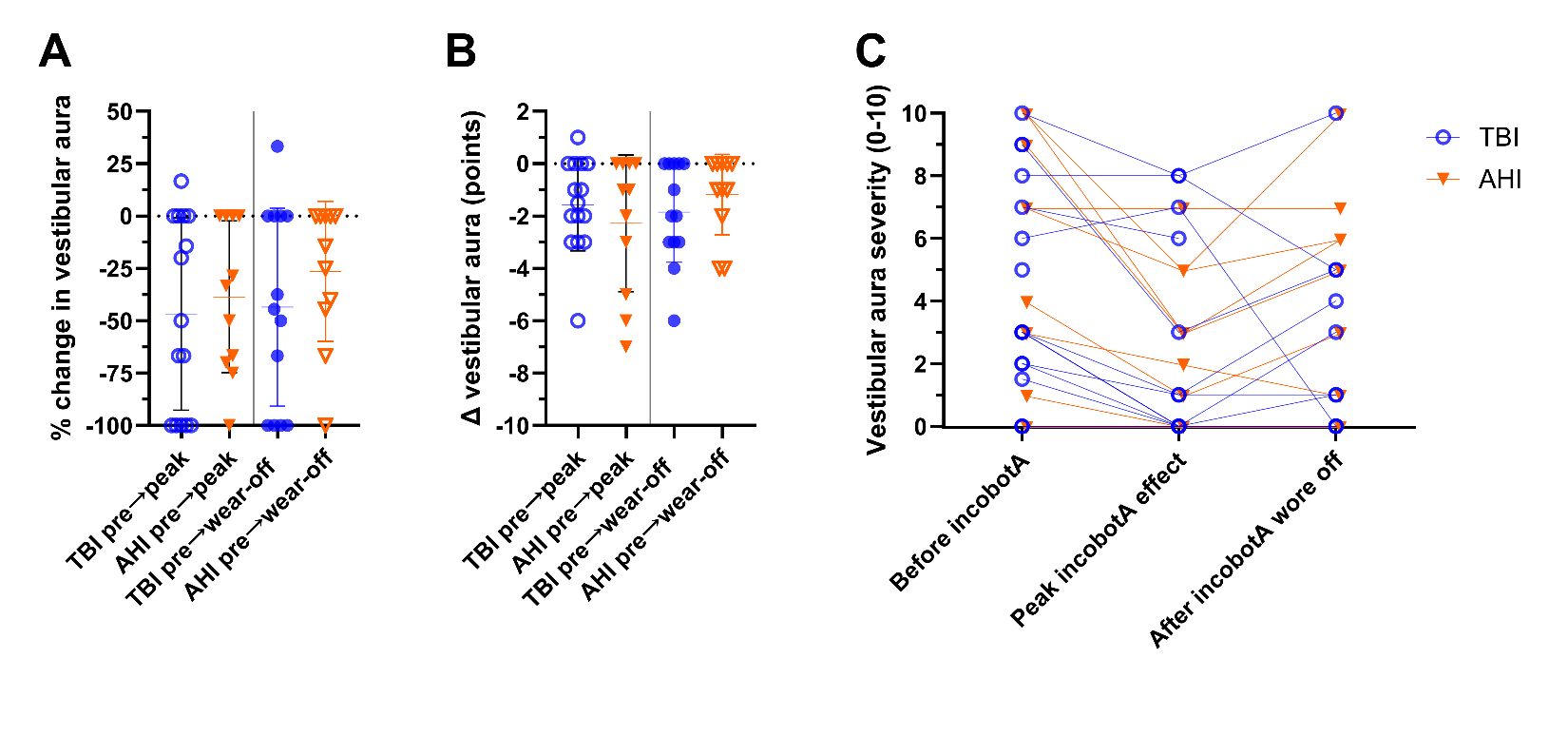

**Supplemental Figure 10. Effects of incobotulinumtoxinA on vestibular aura.**

Self-reported vestibular aura severity is shown before incobotulinumtoxinA treatment (incobotA in the figure), approximately 4 weeks after treatment when peak benefit was expected, and approximately 12 weeks after treatment when treatment effect had typically worn off for participants with traumatic brain injury (TBI; blue circles) and anomalous health incidents (AHI; orange triangles). Panels show percent change in vestibular aura severity (A), absolute change in vestibular aura severity (B), and individual paired severity ratings across timepoints (C). Vestibular aura severity was rated from 0–10. Individual points represent participants; horizontal bars indicate median and interquartile range. Vestibular aura severity decreased at peak treatment effect in both groups, with median reductions of 50.0% in TBI and 33.3% in AHI, but the between-group difference was not significant (p = 0.706). Percent change in vestibular aura severity did not significantly differ between groups from baseline to approximately 12 weeks after treatment (TBI median −44.4%, n = 13; AHI median −14.3%, n = 11; p = 0.402). Absolute change also did not significantly differ between groups from baseline to peak treatment effect (p = 0.726) or from baseline to approximately 12 weeks after treatment (p = 0.467). Between-group comparisons were performed using two-tailed Mann–Whitney U tests.

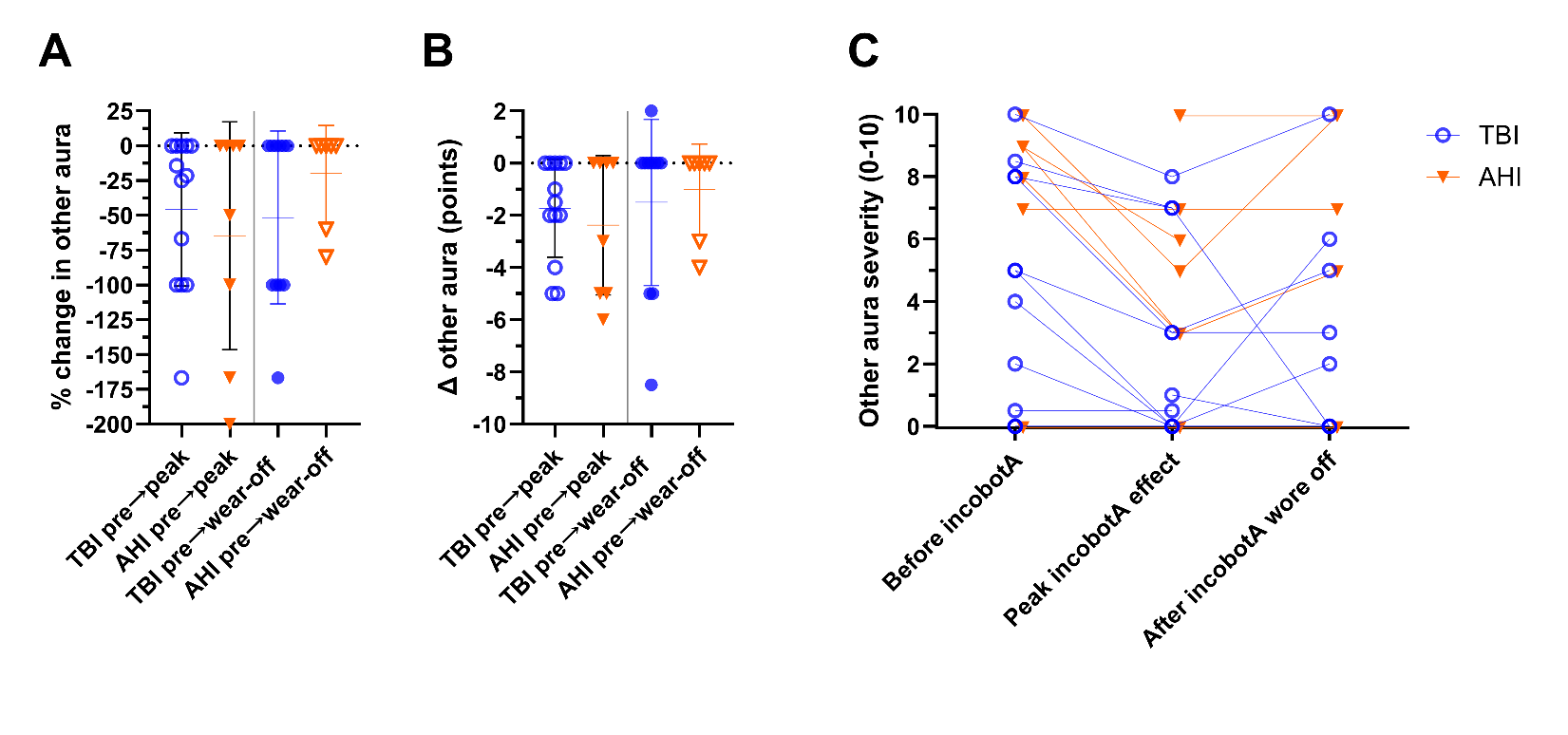

**Supplemental Figure 11. Effects of incobotulinumtoxinA on other aura.**

Self-reported other aura severity is shown before incobotulinumtoxinA treatment (incobotA in the figure), approximately 4 weeks after treatment when peak benefit was expected, and approximately 12 weeks after treatment when treatment effect had typically worn off for participants with traumatic brain injury (TBI; blue circles) and anomalous health incidents (AHI; orange triangles). Panels show percent change in other aura severity (A), absolute change in other aura severity (B), and individual paired severity ratings across timepoints (C). Other aura severity was rated from 0–10. Individual points represent participants; horizontal bars indicate median and interquartile range. Other aura severity decreased at peak treatment effect in both groups, with median reductions of 21.4% in TBI and 25.0% in AHI, but the between-group difference was not significant (p = 0.896). Percent change in other aura severity did not significantly differ between groups from baseline to approximately 12 weeks after treatment (TBI median 0.0%, n = 11; AHI median 0.0%, n = 7; p = 0.253). Absolute change also did not significantly differ between groups from baseline to peak treatment effect (p = 0.741) or from baseline to approximately 12 weeks after treatment (p > 0.999). Between-group comparisons were performed using two-tailed Mann–Whitney U tests.

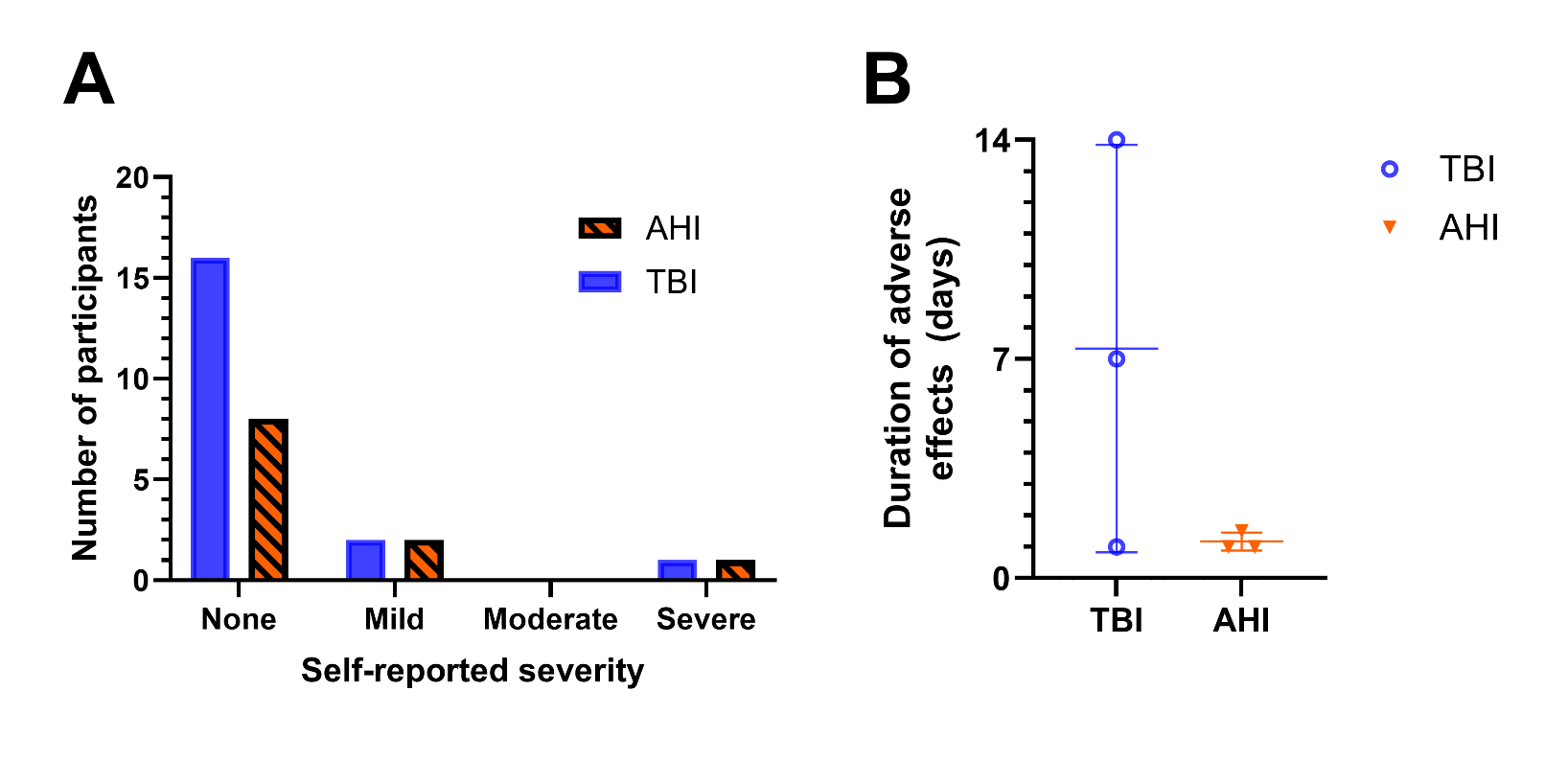

**Supplemental Figure 12. Adverse effects of incobotulinumtoxinA treatments.**

Self-reported adverse effects after incobotulinumtoxinA treatment are shown for participants with traumatic brain injury (TBI; blue) and anomalous health incidents (AHI; orange). Panel A shows the number of participants reporting no, mild, moderate, or severe adverse effects. Severity categories reflected participant self-report and were not independently adjudicated as formal adverse-event severity grades. Most participants in both groups reported no adverse effects. One participant with TBI and one participant with AHI reported a severe adverse effect, while two participants with TBI and two participants with AHI reported a mild adverse effect. Reported adverse effects included facial weakness, eye closure weakness, swelling, and persistent pain; no serious adverse effects requiring hospitalization or discontinuation of incobotulinumtoxinA treatment were reported. Panel B shows the duration of adverse effects in days among participants who reported adverse effects. Individual points represent participants; horizontal bars indicate median and interquartile range. The distribution of adverse effect severity did not significantly differ between groups by Fisher’s exact test (p = 0.809). Duration of adverse effects also did not significantly differ between groups by exact two-tailed Mann–Whitney U test (TBI median 7.0 days, n = 3; AHI median 1.0 day, n = 3; U = 2, p = 0.400).

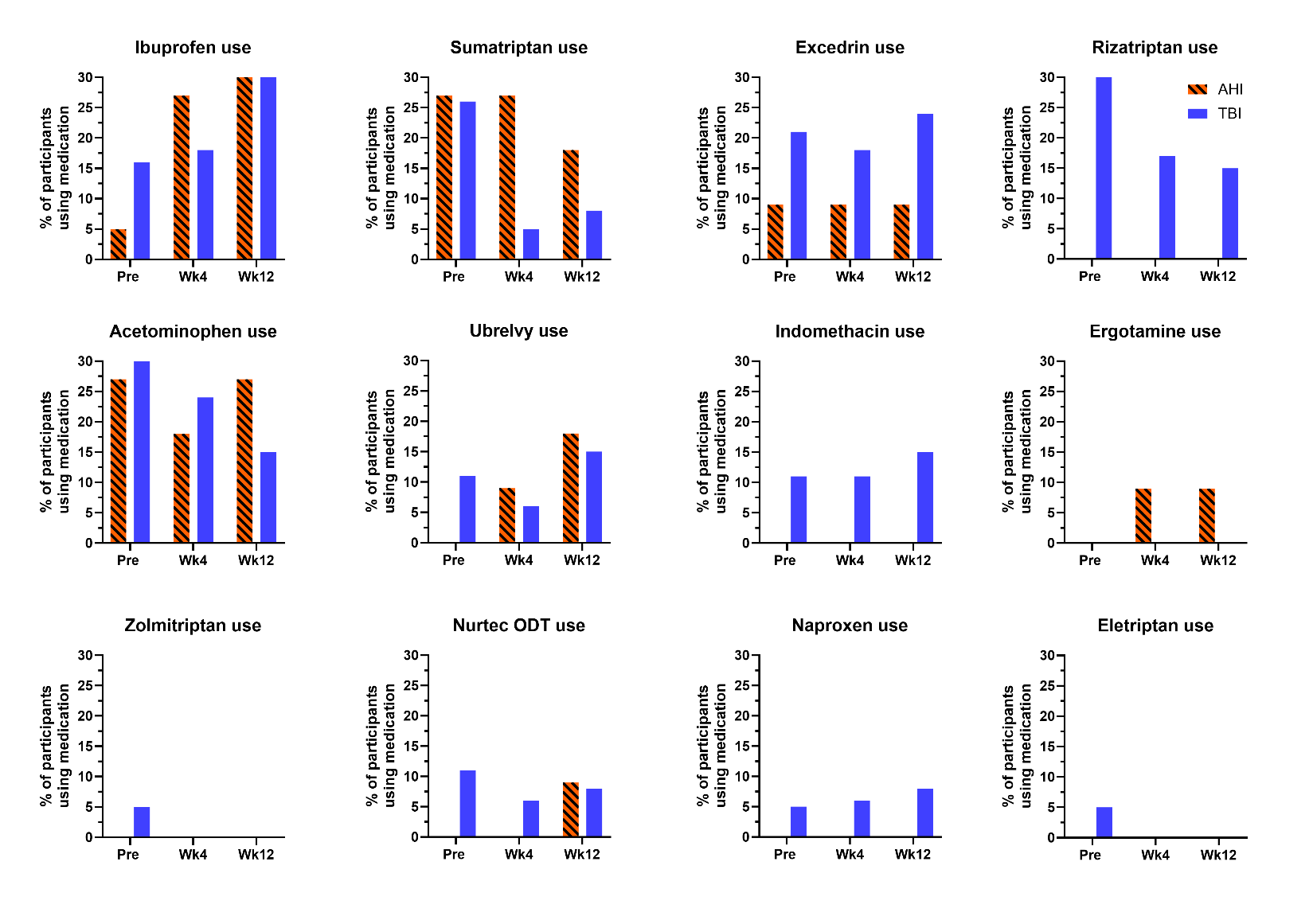

**Supplemental Figure 13. Migraine abortive medications reported.**

Self-reported use of acute abortive migraine medications is shown for participants with anomalous health incidents (AHI; orange hatched bars) and traumatic brain injury (TBI; blue bars) before incobotulinumtoxinA treatment, approximately 4 weeks after treatment, and approximately 12 weeks after treatment. Bars indicate the percentage of participants in each group reporting use of each medication at each timepoint. Participants could report use of more than one medication; therefore, percentages do not sum to 100%. Medication dose, frequency of use, and number of uses per time interval were not systematically captured. Several abortive medications were commonly reported, including NSAIDs or NSAID-containing products, particularly ibuprofen and Excedrin; triptans, particularly sumatriptan; and acetaminophen. Data are presented descriptively; no statistical comparisons were performed.

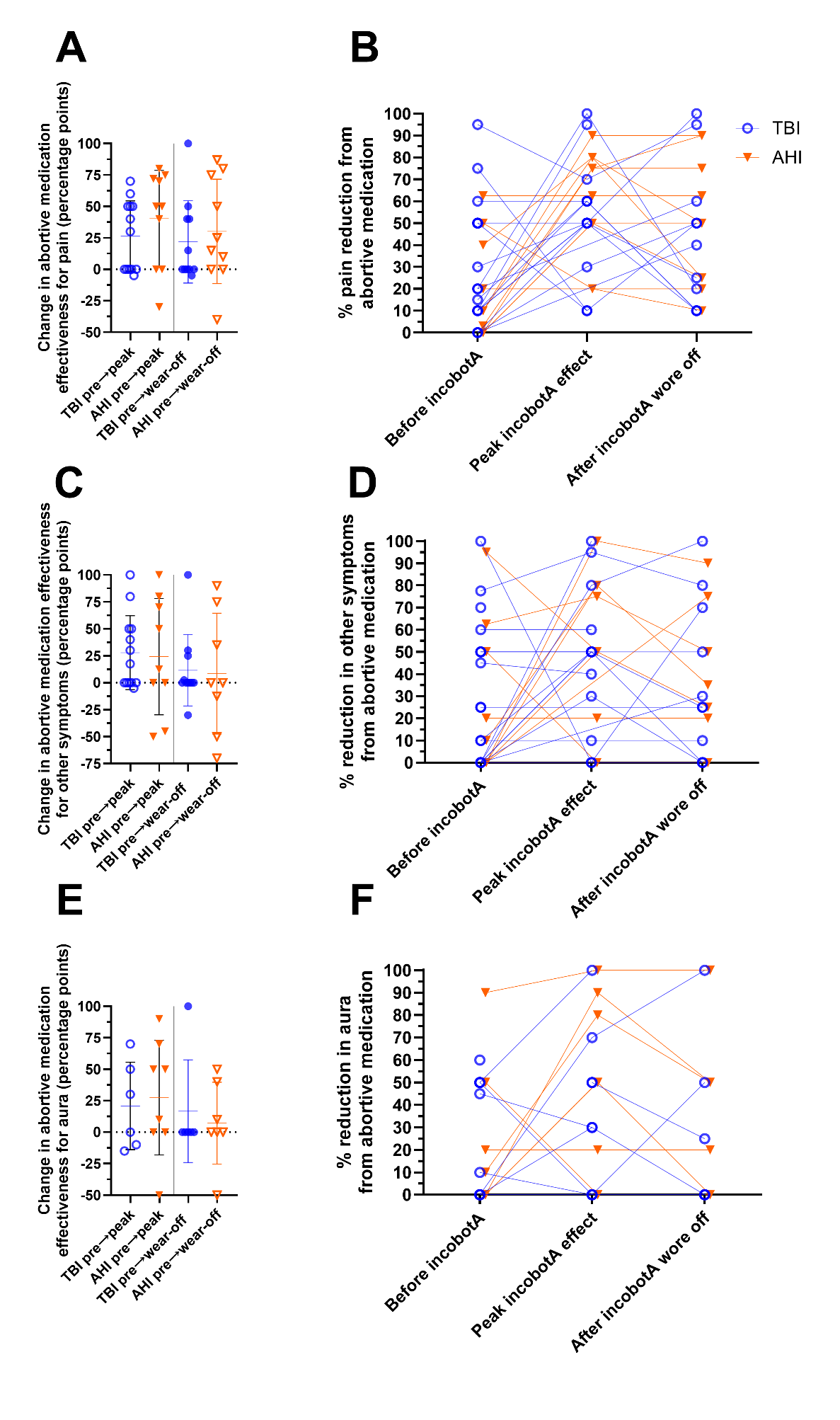

**Supplemental Figure 14. Effectiveness of abortive medications**.

Self-reported effectiveness of acute abortive migraine medications is shown for participants with traumatic brain injury (TBI; blue circles) and anomalous health incidents (AHI; orange triangles) before incobotulinumtoxinA treatment (incobotA in the figure), at peak incobotulinumtoxinA effect, and after incobotulinumtoxinA wore off. Abortive medication effectiveness was rated from 0%–100% for pain relief, reduction in other migraine symptoms, and reduction in aura. The left column shows the change in effectiveness, expressed in percentage points, from before treatment to peak incobotulinumtoxinA effect and from before treatment to after incobotulinumtoxinA wore off for pain relief (A), reduction in other migraine symptoms (C), and reduction in aura (E). The right column shows individual paired ratings across the three timepoints for percent pain reduction from abortive medication(s) (B), percent reduction in other symptoms from abortive medication(s) (D), and percent reduction in aura from abortive medication(s) (F). Individual points represent participants; horizontal bars and error bars in panels A, C, and E indicate the median and interquartile range. Responses were variable across domains, groups, and timepoints. Data are presented descriptively; no statistical comparisons were performed.

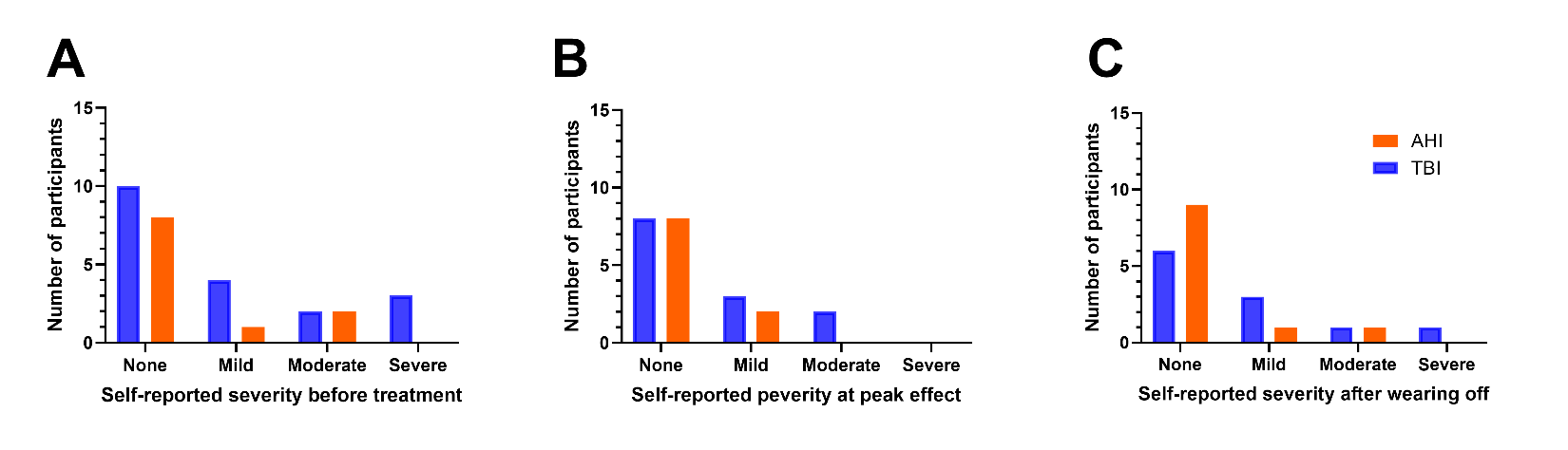

**Supplemental Figure 15. Side effects of abortive medications.**

Self-reported side effects from acute abortive migraine medications are shown for participants with anomalous health incidents (AHI; orange bars) and traumatic brain injury (TBI; blue bars). Panels show the number of participants reporting no, mild, moderate, or severe side effects before incobotulinumtoxinA treatment (A), approximately 4 weeks after treatment (B), and approximately 12 weeks after treatment (C). Most participants in both groups reported no or mild side effects across timepoints, although moderate or severe side effects were reported by some participants. Data are presented descriptively; no statistical comparisons were performed.

**SUPPLEMENTAL DISCUSSION**

This Supplemental Discussion provides additional context for interpretation of the findings, comparison with prior studies, and future research directions.

**Additional clinical observations and study context**

A novel aspect of the study was that we collected self-reported information not just about the presence or absence of migraine features like nausea, phobias, and auras, but also about the intensity of these features on a 0–10 scale. Anecdotally, two patients who were asked to participate but declined were known to have been non-responders.

Anecdotally, self-reported responses to standard treatments for new-onset obstructive sleep apnea, restless leg syndrome, insomnia, attention deficit, and mental fatigue have also appeared similarly effective in patients with AHI vs. patients with TBI. Ongoing and future comparative case series-based studies will be required to document responses in a standardized fashion.

**Mechanistic considerations**

It is possible that many different proximal triggers lead to similar chronic conditions. Since the mechanisms underlying the effects of botulinum toxins on migraine and migraine-like headache disorders are also incompletely understood, it remains possible that while the self-reported effects of incobotulinumtoxinA treatments were similar in the two groups, the mechanisms related to efficacy could still be distinct. To our knowledge, at this time there are no valid animal models of AHI that could be used to investigate these pathophysiological and pharmacodynamic mechanisms in comparison with animal models of TBI-induced migraine-like phenomena.^14,19,23^ Thus, there remain many open questions in this field.

**Comparison with prior literature**

Clinical outcomes were reported to be “better” in 64% of military service members with history of TBI after treatment with onabotulinum toxin A.^12^ In addition, a randomized, placebo-controlled crossover study in military veterans with post-traumatic headache found that botulinum toxin type A significantly reduced the number of headaches per week, headache days per week, and headache pain severity compared with placebo.^13^ A recent scoping review of service members and veterans similarly concluded that botulinum toxin appears promising for migraine and post-traumatic headache in this population, while emphasizing that the evidence base remains limited and under-studied.^24^ A 2022 review of botulinum toxin A treatment in idiopathic migraine (not related to TBI or AHI) reported a range of positive effects in headache frequency, intensity, duration, related disability, and quality of life.^25^ A 2019 meta-analysis reported reduction of 8-9 headache days per month in participants with migraine treated with botulinum toxin A and 5-6 days per month in placebo for a difference of 3 days per month.^26^ This result is very similar to the median reduction of 2 days per week = approximately 8 days per month reported here. It is not known whether botulinum toxin treatments are more effective than other prophylactics. The reduction in headache frequency reported in this study (-67%) is numerically higher than that previously reported for military service members with TBI (-12% for tricyclic antidepressants, -23% for topiramate and -17% for propranolol).^9^ However, the studies were designed differently and are not directly comparable.

**Future directions**

As more information is learned about AHI, it is possible that more specific, mechanism-based interventions may emerge. Future directions for this line of investigation include many further clinical and mechanistic studies. A top priority will be to perform systematic assessments of treatments for new-onset obstructive sleep apnea, restless leg syndrome, insomnia, attention deficit, mental fatigue, vestibular dysfunction, balance impairments, and mood disorders in the contexts of both TBI and AHI. There can be variability from site to site and provider to provider in how treatments are delivered, so broad-based studies will be beneficial. Once there is a clearer understanding of the areas of equipoise and most important unmet needs, then randomized controlled trials should be performed. The status of mechanistic investigations regarding AHI has not been publicly disclosed, so it is not possible to speculate on when, if ever, specific mechanism-based therapeutics will be available. Thus, in the interim, future studies of empirical treatments should be performed to help guide providers caring for patients with AHI.
